# Molecular Diagnosis of Heart Allograft Rejection Using Intra-Graft Targeted Gene Expression Profiling: the heart HistoMX system

**DOI:** 10.1101/2024.09.24.24314238

**Authors:** Alessia Giarraputo, Guillaume Coutance, Jignesh K. Patel, Marny Fedrigo, Olivier Aubert, Jessy Dagobert, Fariza Mezine, Blaise Robin, Philippe Rouvier, Shaida Varnous, Jean Paul Duong Van Huyen, Patrick Bruneval, Annalisa Angelini, Jon Kobashigawa, Alexandre Loupy

## Abstract

**Background and aims:** Tissular gene expression profiling has the potential to refine the diagnosis of cardiac allograft rejection. Contrary to whole-transcriptome approaches, targeted molecular profiling applicable to formalin-fixed paraffin-embedded (FFPE) endomyocardial biopsies (EMB) can be easily implemented in clinical practice. We aimed to develop and validate the first rejection molecular diagnostic system dedicated to heart transplantation (HTx).

**Methods:** An international multicenter study was designed, building a deep phenotyped cohort of HTx recipients recruited between 2011 and 2021 at 4 referral centers. Detailed donors, recipients, clinical, immunological, biological, and histological parameters were collected. EMBs were graded according to international working formulations. Tissue gene expression was analyzed on FFPE-EMB using the consensus Banff Human Organ Transplant gene set. Molecular classifiers of antibody-mediated (AMR) and acute cellular rejection (ACR) were built. Discrimination and calibration were assessed in the development and validation sets (NCT06436027).

**Results:** A total of 591 biopsies were included: 188 AMR (pAMR1(I+): n=51; pAMR1(H+): n=58; pAMR2-3: n=79), 289 ACR (1R n=174; 2-3R n=115) and 114 matched non-rejection cases. Biopsies were split in a derivation (n=476) and a validation set (n=115). AMR top significant transcripts were related to the IFN-gamma inducible pathway, endothelial activation, and monocyte-macrophage recruitment. ACR was characterized by transcripts related to T-cell receptor, CD3 receptor activation, and CD28 signaling. ACR and AMR molecular rejection models were strongly associated with the pathology severity of rejection and accurately identified rejection in the derivation (ROC-AUC: AMR=0.831, ACR:=0.837) and validation sets (ROC-AUC: AMR=0.812; ACR=0.849). Calibration was adequate. The robustness of the molecular classifiers were reinforced by various sensitivity analyzes. An automated report was developed to enhance the reproducibility and clinical applicability of the molecular analysis.

**Conclusions:** In this study, the first tissue-based rejection molecular diagnostic system applicable to FFPE- EMB and dedicated to heart transplantation rejection was developed and internally validated. This tool has the potential to refine the diagnosis of rejection.

## INTRODUCTION

Heart transplantation (HTx) remains the most valuable therapeutic option for patients with end-stage heart failure refractory to optimal medical therapy (PMID: 27206819, 28455343). More than 6,000 heart transplants are performed annually worldwide (PMID: 34419370) and the median post-transplant survival now exceeds 12.5 years.

Despite major improvements in immunosuppression and transplant care, acute and chronic rejection-induced allograft injuries remain one of the leading causes of mortality and morbidity after heart transplantation, thus limiting recipients’ life expectancy. An improvement in the overall management of rejection remains an unmet medical need (PMID: 30776902, 29452978). As a first step, a precise diagnosis of rejection is crucial to guide patient care and optimize management. Nowadays, the diagnosis of cardiac rejection still relies exclusively on the pathological assessment of endomyocardial biopsies (EMBs) by identifying and grading cellular infiltrates and myocardial damage (PMID 24263017, 21555100).

While important advances have been made in the standardization of the rejection diagnosis, pathology remains an imperfect gold-standard, particularly due to the important inter-observer variability (PMID: 23222738), sample bias and the use of qualitative or semi-quantitative scales that oversimplify complex phenotypes (PMID: 34954735). Additionally, disease severity, degree of myocardial injury and progression stage are crucial pieces of information poorly captured by the current working formulations. All these limits represent major barriers to achieve a precise and reliable diagnosis of rejection. In this context, gene expression profiling of fresh myocardial samples and using a whole transcriptome approach arose as a potential objective companion tool of pathology to refine the diagnosis of rejection (PMID 29178397, 29178397, 28662985). However, important drawbacks have limited the routine clinical applicability of whole-transcriptome based diagnosis including extra-core sampling bias, low reproducibility, technical and analytical heaviness, costs and sample turn-around that is not compatible with a clinical setting. Moreover, the lack of access to archival samples for the development and validation of models is another key issue that may explain why previous cardiac allograft rejection molecular classifiers have been developed on kidney biopsies and not EMB (PMID: 28662985).

Recent technologies, such as the Nanostring nCounter system, could overcome this limitation by allowing analysis of the same tissue used for histology (PMID: 26069246). Targeted molecular profiling applicable to formalin-fixed paraffin-embedded (FFPE) endomyocardial biopsies may allow the implementation of molecular diagnosis into the clinical routine. Recently, we have shown that the Banff Human Organ Transplant Panel (B-HOT) panel, a consortium-approved panel comprising 770 genes, developed by the Banff Molecular Diagnostics Working Group (MDWG) (PMID: 32428337), accurately captured key molecular patterns of antibody-mediated rejection (AMR) in heart allograft biopsies and may serve as a proxy to whole transcriptome-based analysis (PMID: 37745639).

The aim of the present study was to identify gene expression signatures for AMR and acute cellular rejection (ACR) in heart transplantation and to develop and validate a molecular heart rejection diagnostic system based on targeted transcriptome as a novel monitoring companion tool for heart allograft precision diagnostics applicable to routine clinical practice (**Figure 1**).

**Figure 1.**
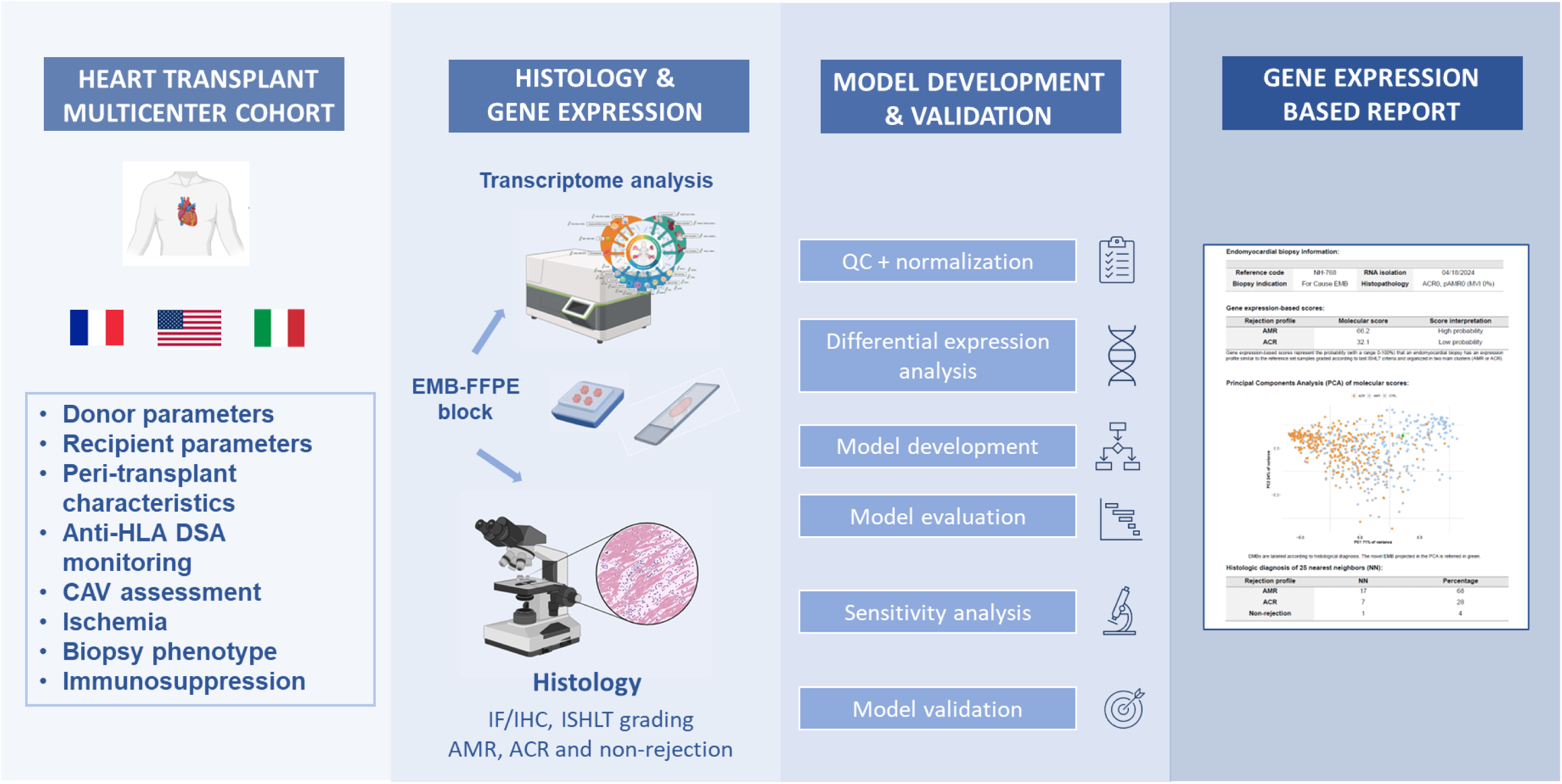
Study design. We developed a deeply phenotyped cohort of heart transplant recipients from 3 international referral centers. Endomyocardial biopsies were collected and sequenced for transcriptome profiling. Tissue-based molecular models were developed and validated externally. An automated gene expression-based report was implemented to apply rejection models on novel biopsies.

## METHODS

### Study design, patients and endomyocardial biopsies selection

We designed an international multicenter study, building a deep phenotyped cohort of heart transplant recipients recruited at 4 referral centers (Pitié-Salpêtrière and Georges Pompidou hospitals in Paris, University General Hospital of Padova and Cedars Sinai Medical Center in Los Angeles, USA). FFPE-EMBs were collected as part of routine clinical care between 2011 and 2021 and graded according to the ISHLT working formulations (PMID: 16297770, PMID: 24263017). Biopsies were selected according to predefined criteria (Supplementary Methods) to be representative of the spectrum of cardiac allograft rejection. Ambiguous cases including biopsy-negative rejection, mixed rejection, and Quilty cases were excluded. Control biopsies were matched with rejection cases for transplant year, time post-transplant and recipient gender.

All patients were included following written informed consent for a protocol approved by the local institutional review boards (Comité de Protection des Personnes Île de France II – research project 20141226, Institutional Review Board registration 00001072; Ethics committee Azienda-Hospital University Padua, protocol number 0062556/2022; Institutional Review Board of Cedars-Sinai Medical Center). All procedures were carried out according to the Declaration of Helsinki as well as the ethical standards of the institutional and national research committee.

### Data collection

A comprehensive patients and biopsies data collection has been performed at all participating centers in a centralized and anonymized database to ensure harmonization across study centers. Data collected were related to (i) recipients (age, sex, cause of heart failure) and donors baseline characteristics (age, sex), (ii) transplant procedure parameters (ischemic time, presence of severe primary graft dysfunction), (iii) immunosuppressive therapy (induction therapy, maintenance immunosuppressive regimen), (iv) clinical (indication of EMB, time post-transplant, presence of symptoms or signs of heart failure), (v) immunological (circulating anti-HLA donor-specific antibodies), (vi) echocardiographic (left ventricular ejection fraction) and (vii) histological parameters (ACR and AMR rejection grades according to ISHLT working formulations).

### Procedures and clinical protocols

#### Histology and immunohistochemistry

EMB were performed for routine clinical purposes (either protocol or for-cause EMB) through jugular or femoral access and were formalin-fixed, paraffin-embedded, and routinely stained with hematoxylin and eosin (HE) for histopathologic evaluation, processed and evaluated following international standards (PMID: 20643330, PMID: 37080658). All EMB were graded by expert pathologists according to the latest international working formulations (PMID: 16297770, PMID: 24263017). Immunostaining was performed on tissue sections with specific antibodies, as previously described (PMID: 25612500, **Supplementary material**). C4d staining was performed on all biopsies. Grading for C4d staining was performed by scoring pattern and intensities in a semi-quantitative way with a grade range from 0 to 3 following the working formulation of ISHLT (PMID: 21555100). Only C4d positive EMB were considered for the diagnosis of pAMR1(I+) cases. Mixed rejection was defined by the association of ACR ≥ 2R and AMR ≥ pAMR2. All EMBs were read by two highly-trained cardiovascular pathologists (PB, PR, MF, AA). Disagreements were solved by a consensus.

#### Donor specific antibodies

Anti-HLA donor-specific antibodies (DSA) were evaluated by Luminex technology (Luminex LABScreen Single Antigen, One-Lambda Inc., West Hills, CA, USA) as previously described. A mean fluorescence intensity (MFI) value of 1000 was considered positive (PMID: 23763485).

We defined “immunodominant DSA” as the DSA with the highest MFI, either class I or II. Only tests performed at the time of the EMB or (i) in the previous 3 months for EMB performed during the first-year post-transplant or (ii) in the previous 6 months for EMB performed beyond the first-year post-transplant were considered.

#### Outcomes measure

The reference diagnosis of rejection implemented in the model development was based on the histopathological assessment of cardiac allograft biopsies. Diagnostic categories were referred to as antibody-mediated- and acute cellular rejection as well as no evidence of rejection event. For all molecular analyses, ACR was defined as an ACR ≥ ACR 1R and AMR as AMR ≥ pAMR1 (PMID: 16297770, PMID: 24263017).

The primary outcome was the assessment of the prediction performances of the molecular classifiers (ACR and AMR rejection scores) in the internal validation set using a multiparametric approach to predict biopsy-proven rejection defined as ACR ≥ ACR 1R and AMR ≥ pAMR1 for the ACR and AMR molecular models, respectively.

#### Sample size calculation

To ensure the robustness and reliability of our findings, a comprehensive sample size calculation was performed. The minimal sample size was determined based on an acceptable difference of 0.05 between the adjusted and apparent R-squared values. This difference reflects the anticipated level of precision in our statistical model. A margin of error of 0.05 was deemed acceptable, to balance the risk of Type I and Type II errors. Additionally, a desired statistical power of at least 0.8 was set, meaning there is an 80% probability of detecting a true event of rejection. This power threshold helps to mitigate the risk of Type II errors, thereby increasing the likelihood that the study findings will be both valid and replicable. Incorporating these parameters into our calculations, the required minimum sample size was determined to be 400 biopsies in the derivation set. This substantial sample size was necessary to ensure the statistical robustness of our molecular approach, allowing for reliable detection of differences in immune cell infiltration, gene expression signatures.

### Gene expression profiling

The overall pipeline of the molecular analysis from the initial FFPE biopsy to the complete analysis report is described in **Supplementary Figure S1**.

#### RNA isolation from formalin-fixed paraffin-embedded biopsies

To obtain 100 ng of RNA required for the transcriptome sequencing, three 20-µm thick sections were cut from each paraffin block. The RNA isolation process comprehended three key steps: i) deparaffinization, ii) removal of contaminant (both organic and DNA related); iii) final serial centrifugation to remove salt contaminant and concentrate isolated RNA (**Supplementary Methods** for full procedure). Total RNA was isolated using the Rneasy FFPE Kit (Qiagen, Hilden, Germany). A quantity and quality assessment of RNA was performed before downstream steps using the Nanodrop 2000 spectrophotometer (Thermo Fisher Scientific Inc., Waltham, MA, USA).

#### Transcriptome sequencing

Gene expression analysis was performed with the nCounter technology (Nanostring) using the targeted gene Banff Human Organ Transplant Panel (B-HOT panel) (PMID: 32428337). This panel was designed by the Banff Molecular Diagnostics Working Group and covers 770 genes (758 endogenous and 12 housekeeping genes) representative of the key functional pathways involved in solid organ transplantation rejection. The nCounter technology relies on the digital detection and direct molecular barcoding of targeted mRNA through the design of two specific probes (capture and reporter probes) for each mRNA (PMID: 18278033). This direct assessment has the major asset to bypass amplification steps, thus reducing technical variations and probe instability.

### Bioinformatic and statistical analysis

#### Quality control and normalization

Gene expression data raw counts were analyzed with the nSolver Analysis Software (version 4.0.70) and subjected to extensive quality control metrics, related to: (i) imaging, (ii) binding density, (iii) linearity of the positive controls and iv) and limit of detection (**Supplementary Methods**). Samples that fail to meet at least one of the quality control metrics were flagged and then excluded from further analysis. Pre-normalization steps with assessment of housekeeping genes stability, and data visualization to detect potential batch effects was performed (**Supplementary Methods**). Raw gene expression data were normalized using the Reduced Unwanted Variation method (PMID: 22513995, PMID: 32789507, **Supplementary Methods**). Overall, quality control, housekeeping genes stability and normalization steps allowed us to correct the sequencing data for technical variations. A total of 610 EMBs from 461 patients were analyzed, including 192 antibody-mediated rejection, 296 acute cellular rejection and 122 non-rejection biopsies. Among them, 19 (3.1%) biopsies did not meet quality control criteria and were excluded from further analyzes.

Biopsies validating all these pre-analytical steps were included in the final molecular reference set. Overall, the cohort consisted of 591 biopsies from 455 patients, comprising 188 AMR cases, 289 ACR cases and 114 non-rejection cases, which were then divided into a training and a validation set with 80:20 proportion using the *dataPartition* function of caret package to homogeneously stratify rejections events across the training and validation sets (training set: 152 AMR, 232 ACR, 92 non-rejection cases; validation set: 37 AMR, 57 ACR, 22 non-rejection cases).

#### Differential expression and pathway analysis

Differential expression analysis was conducted using the *DESeq* function (DESeq2 package, PMID: 25516281) applying a Wald test to compare gene expression between the rejection condition of interest versus everything-else. Differentially expressed genes were identified from the linear fitting after calculating fold change for each comparison of interest. P-values were adjusted for multiple comparisons using the Benjamini-Hochberg method for false discovery rate (FDR). Significant genes were filtered with a FDR p-value lower than 0.05. To control false discovery rate, a shrinkage procedure was applied. To annotate the differentially expressed genes, we used Uniprot, Gene Cards database and the Banff Molecular Report reference (PMID: 29425356, 27322403, 32428337). Finally, pathways enrichment analysis was performed to characterize the pathophysiological relation between the most differentially expressed genes (Over Representation Analysis method (PMDI: 15297299), through the *enrichplot* package. A hypergeometric model with Reactome repository was applied. Opportune pathways’ identifiers labeling, and general membership class were retrieved from the repository. All statistical and bioinformatic analyses were performed using R (version 4.0.5 R Foundation for Statistical Computing).

#### Development and validation of molecular classifiers of rejection

Supervised molecular classifiers were then developed in the training set for AMR, ACR and non-rejection and were evaluated in an equally partitioned validation set. The top 20 ranked differentially expressed genes for AMR and ACR were the input variables for the multivariate logistic regression model (*glm* function from caret package). The supervised model was trained using 10 K-fold cross validation (CV) as a resampling procedure. To move from molecular scores to interpretable diagnosis, we applied a threshold derived from the Youden index of the continuous predicted probabilities to classify each case as either AMR, ACR, or non-rejection. Rejection-related classifiers were then evaluated for model performance, using a multiparametric approach: precision-recall area under the curve (PR-AUC), diagnostic accuracy, positive predictive value (PPV), true positive rate (TPR), true negative rate (TNR), receiver operating characteristics (ROC)-AUC, Brier score and F1 score. Confusion matrix was used to display the classification accuracy of the predictive model, comparing the agreement between the histopathological evaluation and the predictive molecular models. Finally, the stepwise increase in rejection-related gene expression with severity of rejection was tested for both AMR and ACR (rejection scores, principal component analysis).

A molecular map of cardiac allograft rejection was represented using a principal component analysis (PCA) based on the individual molecular scores and reflected the molecular heterogeneity of rejection.

#### Sensitivity analyses

Sensitivity analyses were performed to evaluate the robustness of molecular rejection diagnostic models. First, feature selection methods (Maximum Relevance - Minimum Redundancy and random forest models) were applied to optimize gene selection to reduce correlation and collinearity while maintaining similar model performance. Second, EMB for patients with ≥2 biopsies included in the reference set were randomly selected to evaluate the stability of the models when including only one EMB per patient as well as time post-transplant. Third, to minimize the risk of oversight of the minority class (AMR), four balancing methods were applied (Upsampling, Downsampling, SMOTE, ROSE).

## RESULTS

### Characteristics of patients and endomyocardial biopsies

Overall, the cohort consisted of 591 biopsies from 455 patients. Baseline characteristics of recipients, donors and transplant procedures are detailed in **Table 1**. Mean recipient age was 51.1±13.05 years old. Patients were mostly males with the main cause of heart failure related to dilated cardiomyopathy. Routine immunosuppression was mostly based on a combination of calcineurin inhibitors, mycophenolate mofetil and corticosteroids (n = 389, 85.4%).

**Table 1.**
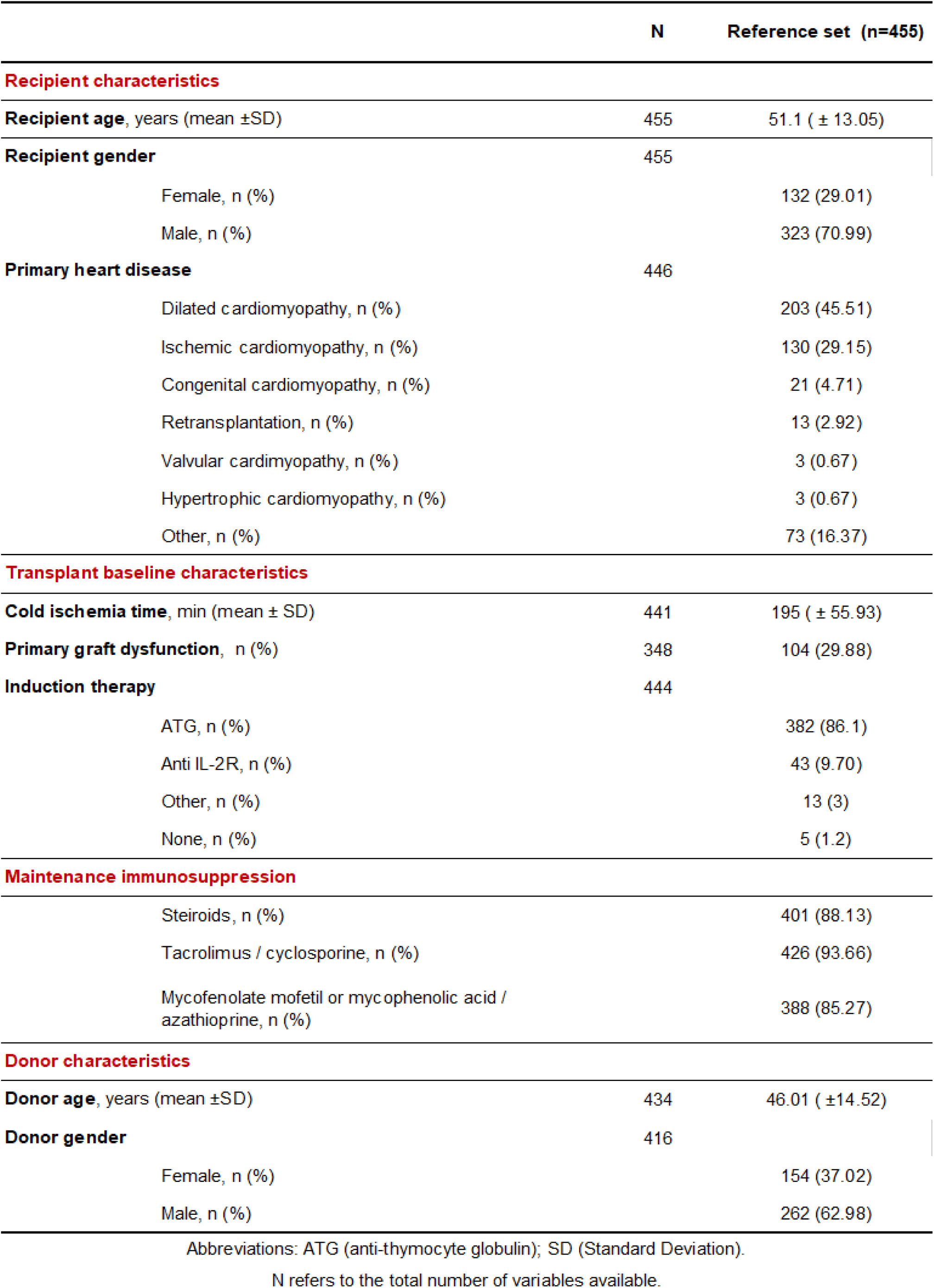
Baseline characteristics of patients (n=455)

|  | **N** | **Reference set (n=455)** |
| --- | --- | --- |
| **Recipient characteristics** |  |  |
| **Recipient age**, years (mean ±SD) | 455 | 51.1 ( ± 13.05) |
| **Recipient gender** | 455 |  |
| Female, n (%) |  | 132 (29.01) |
| Male, n (%) |  | 323 (70.99) |
| **Primary heart disease** | 446 |  |
| Dilated cardiomyopathy, n (%) |  | 203 (45.51) |
| Ischemic cardiomyopathy, n (%) |  | 130 (29.15) |
| Congenital cardiomyopathy, n (%) |  | 21 (4.71) |
| Retransplantation, n (%) |  | 13 (2.92) |
| Valvular cardiomyopathy, n (%) |  | 3 (0.67) |
| Hypertrophic cardiomyopathy, n (%) |  | 3 (0.67) |
| Other, n (%) |  | 73 (16.37) |
| **Transplant baseline characteristics** |  |  |
| **Cold ischemia time**, min (mean ± SD) | 441 | 195 ( ± 55.93) |
| **Primary graft dysfunction**, n (%) | 348 | 104 (29.88) |
| **Induction therapy** | 444 |  |
| ATG, n (%) |  | 382 (86.1) |
| Anti IL-2R, n (%) |  | 43 (9.70) |
| Other, n (%) |  | 13 (3) |
| None, n (%) |  | 5 (1.2) |
| **Maintenance immunosuppression** |  |  |
| Steroids, n (%) |  | 401 (88.13) |
| Tacrolimus / cyclosporine, n (%) |  | 426 (93.66) |
| Mycophenolate mofetil or mycophenolic acid / azathioprine, n (%) |  | 388 (85.27) |
| **Donor characteristics** |  |  |
| **Donor age**, years (mean ±SD) | 434 | 46.01 ( ±14.52) |
| **Donor gender** | 416 |  |
| Female, n (%) |  | 154 (37.02) |
| Male, n (%) |  | 262 (62.98) |

Abbreviations: ATG (anti-thymocyte globulin); SD (Standard Deviation).
N refers to the total number of variables available.

Biopsies were representative of the landscape of heart allograft rejection pathology and included 188 AMR cases (pAMR1(I+): n=51; pAMR1(H+): n=58; pAMR2-3: n=79), 289 ACR cases (ACR 1R n=174; ACR 2-3R, n=115) and 114 non-rejection cases (ACR 0, pAMR0). Median time between transplant and EMB was 238 days (IQR 88-844). A total of 60.1% of EMB were performed during the first-year post-transplant. Biopsies were mostly protocol EMB (86%). At the time of biopsy, anti-HLA DSA were detected in 193 (32.65%) cases. An acute allograft dysfunction defined as a left ventricular ejection fraction (LVEF) < 0.45 and/or an absolute drop in ≥ 0.15 was present in 43 (7.3%) cases. Only 17.5% of the patients have evidence of CAV grade 1 from coronarography (Table 2).

**Table 2.** Endomyocardial biopsy characteristics (N=591)

|  | N | Biopsies included (n=591) |
| --- | --- | --- |
| **Characteristics of endomyocardial biopsies** |  |  |
| **EMB before 1-year post-transplant, n (%)** | 591 | 355 (60.1) |
| **Time between HTx and EMB, median (25th–75th percentile)** | 591 | 238 (88–844) |
| **Type of EMB** | 585 |  |
| For Cause, n (%) |  | 79 (13.50) |
| Protocol, n (%) |  | 506 (86.50) |
| **Cardiac function and events** |  |  |
| **ISHLT 2013 pAMR grade** | 591 |  |
| pAMR0, n (%) |  | 403 (68.19) |
| pAMR 1 (H+), n (%) |  | 58 (9.81) |
| pAMR 1 (I+), n (%) |  | 51 (8.63) |
| ≥ pAMR 2, n (%) |  | 79 (13.36) |
| **ISHLT 2004 ACR grade** | 591 |  |
| 0R, n (%) |  | 302 (51.10) |
| 1R, n (%) |  | 174 (29.44) |
| ≥ 2 R, n (%) |  | 115 (19.46) |
| **LVEF at time of EMB, median (25th–75th percentile)** | 540 | 63 (58 - 66) |
| **Graft dysfunction, %** | 540 | 43 (7.3) |
| **Angiography at time of EMB** | 486 |  |
| CAV grade 0, n (%) |  | 401 (82.51) |
| CAV grade 1, n (%) |  | 85 (17.49) |
| **Anti-HLA DSA at the time of index EMB, n (%)** | 385 |  |
| DSA positive, n (%) |  | 201 (52.20) |
| DSA class (I), n (%) |  | 64 (32) |
| DSA class (II), n (%) |  | 136 (68) |

Abbreviations: HTx (heart transplantation); AMR (antibody-mediated rejection); ACR (acute cellular rejection); CAV (cardiac allograft vasculopathy); DSA (donor-specific antibody); EMB (endomyocardial biopsy); ISHLT (International Society for Heart and Lung Transplantation); LVEF (left ventricular ejection fraction).
N refers to the total number of variables available

### Molecular patterns of rejection in heart transplantation

#### Antibody mediated rejection

Analysis of the differentially expressed genes (DEG) comparing AMR cases to everything else (ACR and non-evidence of rejection) is depicted in **Figure 2A**. The top 20 ranked DEG revealed an increased expression of transcripts related to adaptive immune response (TM4SF18), NK-cells (FCGR3A/B), endothelial cells (PECAM1, NOS3, KLF4, CAV1, ROBO4), donor specific antibody (PLAT), IFN-gamma inducible pathway (CXCL11, CX3CL1, PLA1A, CCL19), cytokine signaling (BAFT, IL1R2, LTB), neutrophil degranulation (CLEC4C), Th2 dedifferentiation (GATA3), and monocyte-macrophage recruitment (LILRB1) (**Supplementary Table S1**). The top significant AMR-associated pathways derived from DEG were related to interferon gamma (q= 1.97E-05), interleukin signaling (q=1.63E-07), toll-like receptor (q=7.83E-12) co-stimulation by CD28 (B-cell activation, q=2.23E-03), antigen processing cross-presentation (q=3.76E-04), lymphoid and non-lymphoid immune interaction (q=7.17E- 06), chemokine binding (q=7.83E-12) and NF-kB pathway (q=3.27E-04) (**Supplementary Figure S2**).

**Figure 2.**
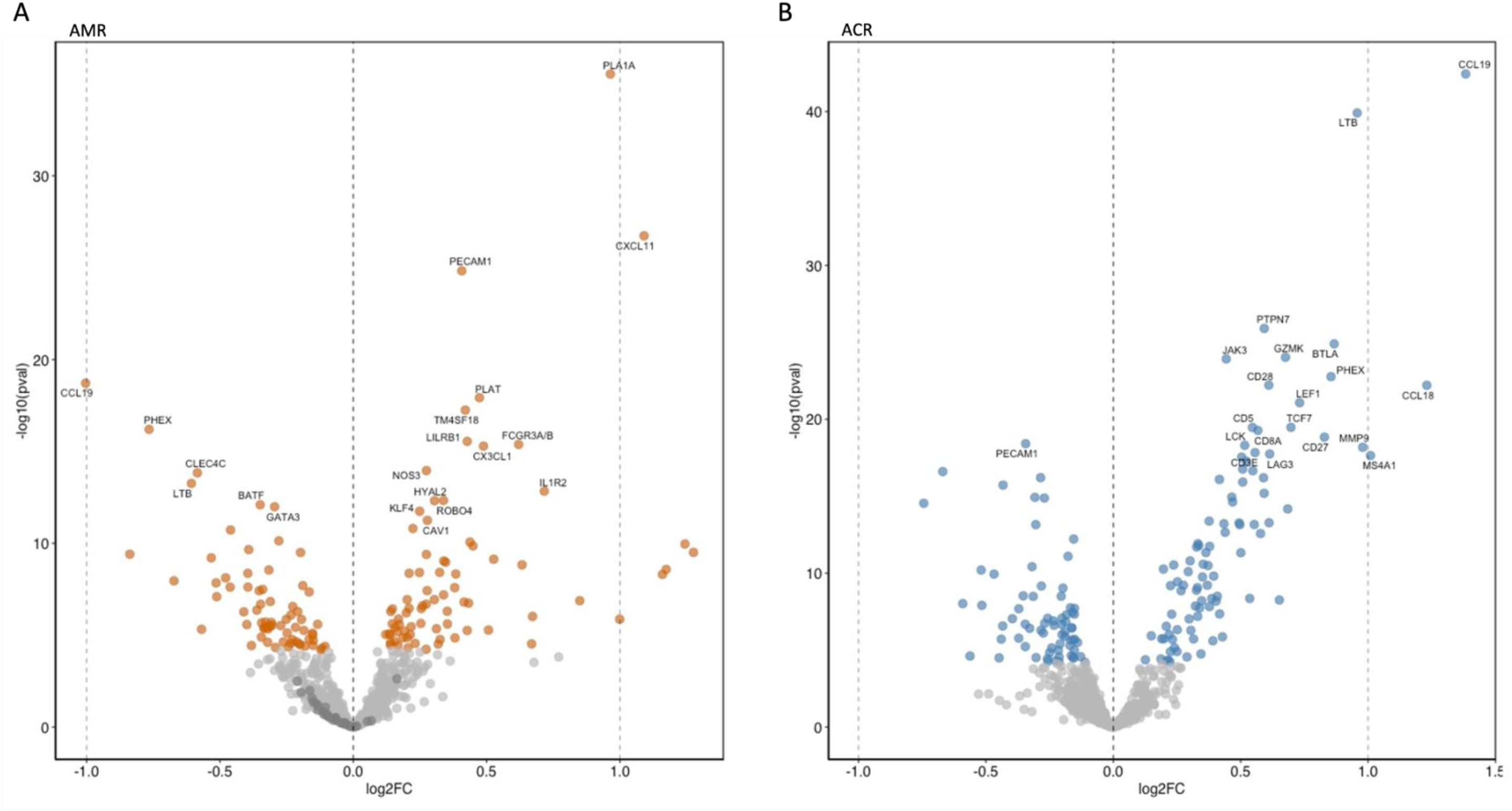
Differential gene expression analysis of antibody-mediated and acute cellular rejection in heart allograft biopsies. Panel. **A.** Volcano plots of differentially expressed genes associated with AMR. The top differentially expressed genes were related to NK-cells, endothelial cells, IFN-inducible pathway, and the monocyte-macrophage recruitment. **Panel B.** Volcano plots of differentially expressed genes associated with ACR showed most significant genes associated with T-cell recruitment, activation of innate and adaptive immune response, CD3 receptor activation, and CD28 signaling. Dots represent individual transcripts. Light blue points indicate genes with a significant p-value adjusted for FDR < 0.05. The x axis illustrates the log2 fold change and on the y axis false discovery rate log 10 transformed for the association of each transcript with rejection profile compared to everything else. ACR, acute cellular rejection; AMR, antibody-mediated rejection.

#### Acute cellular rejection

The top DEG identified were comparing ACR cases to everything else were related to T-cell receptor signaling (CD8A), T-cell checkpoint signaling (CD27, LAG3), natural killer cell and innate lymphoid cell development (TCF7), T-cell mediated rejection (PTPN7), B-cells (MS4A1), IFN-gamma inducible (CCL19), quantitative constitutive macrophage (CCL18), TNF family signaling (LTB, MMP9),CD3 receptor activation (CD3E), chemokine signaling (JAK3), cytotoxicity (GZMK, LCK), and CD28 costimulation signaling (BTLA, CD28) (**Figure 2B, Supplementary Table S2**).

The top significant ACR-associated pathways were related to TCR signaling cascade (q= 5.14E-06), IL-2 signaling (q= 5.14E-06), phosphorylation of CD3 subunit (q= 2.55E-07), cell surface interaction (q= 8.14E-09) and chemokine receptors (q= 8.47E-13) (**Supplementary Figure S3**).

### Development of a gene expression diagnostic system

The derivation set included a total of 475 randomly selected EMB distributed as follows: 151 AMR (pAMR1(I+): n=36; pAMR1(H+): n=46; pAMR2-3: n=69), 232 ACR (ACR 1R n=143; ACR 2-3R, n=89), and 92 non-rejection cases. The 20 most DEG in ACR and AMR were the input variables of the multivariate logistic regression. Using a 10-fold cross validation approach, supervised molecular classifiers were developed and predicted probability for each AMR and ACR model were generated.

AMR and ACR molecular scores were strongly associated with the pathology assessment of severity of rejection according to AMR and ACR international working formulations, respectively (ACR: p for trend = 1.017E-46, AMR: p for trend = 2.911E-52, **Figure 3A**). Molecular AMR score was not associated with ACR severity; neither was the ACR molecular score with AMR severity.

**Figure 3.**
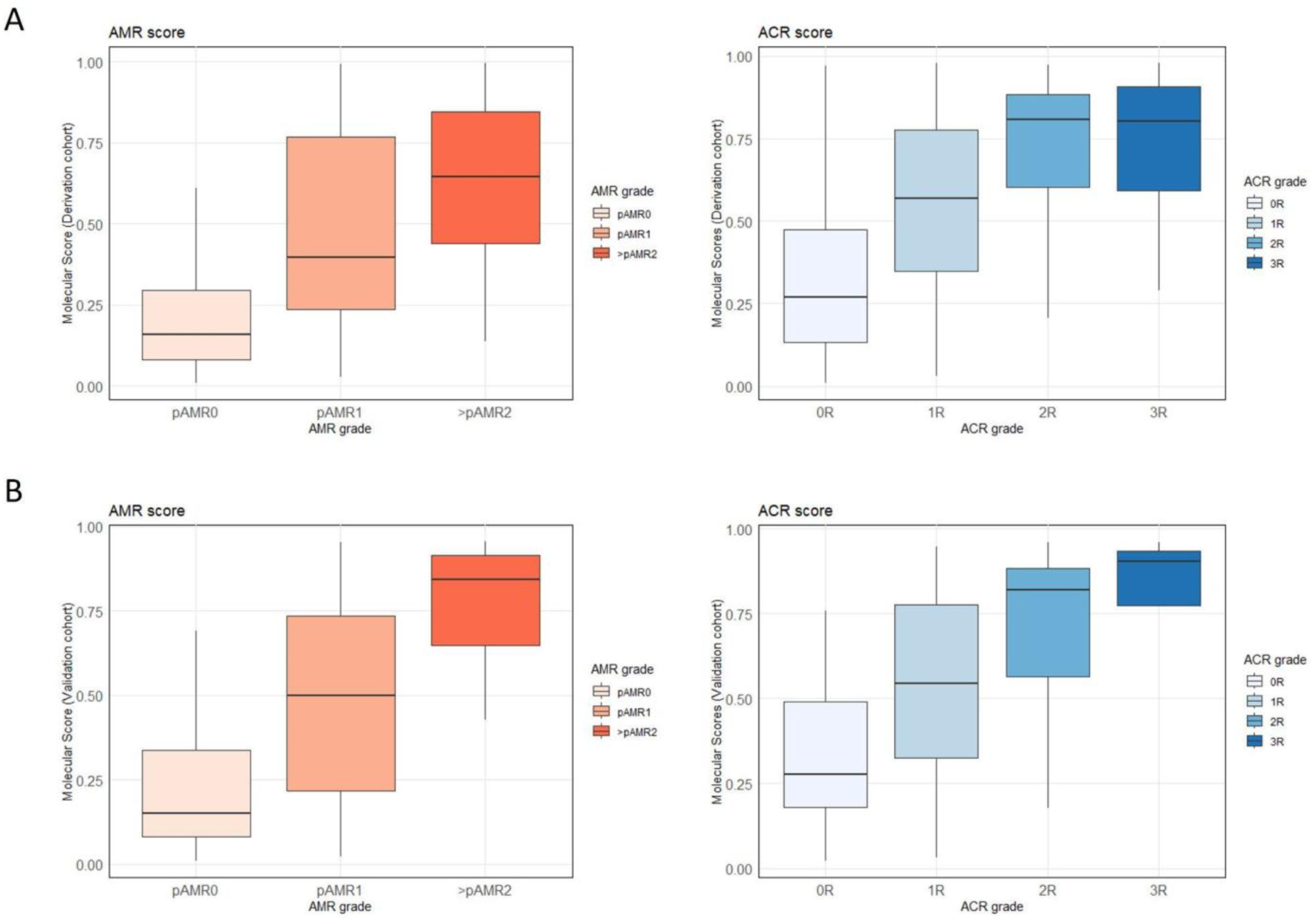
Association between molecular scores and pathology severity of rejection. Panel. **A.** Derivation set. **Panel B**. Validation set. We observed a stepwise increase in AMR and ACR molecular scores with increasing disease severity as assessed with conventional histology (ISHLT working formulations), both in the derivation and the validation cohorts. ACR, acute cellular rejection; AMR, antibody-mediated rejection.

Principal component analysis derived from molecular rejection probability scores was used to visualize the molecular heterogeneity of cardiac allograft rejection. Rejection probabilities derived from the molecular scores discriminated between the two main rejection profiles as well as non-rejection related biopsies (principal component 1: variance 72%; **Figure 4)**.

**Figure 4.**
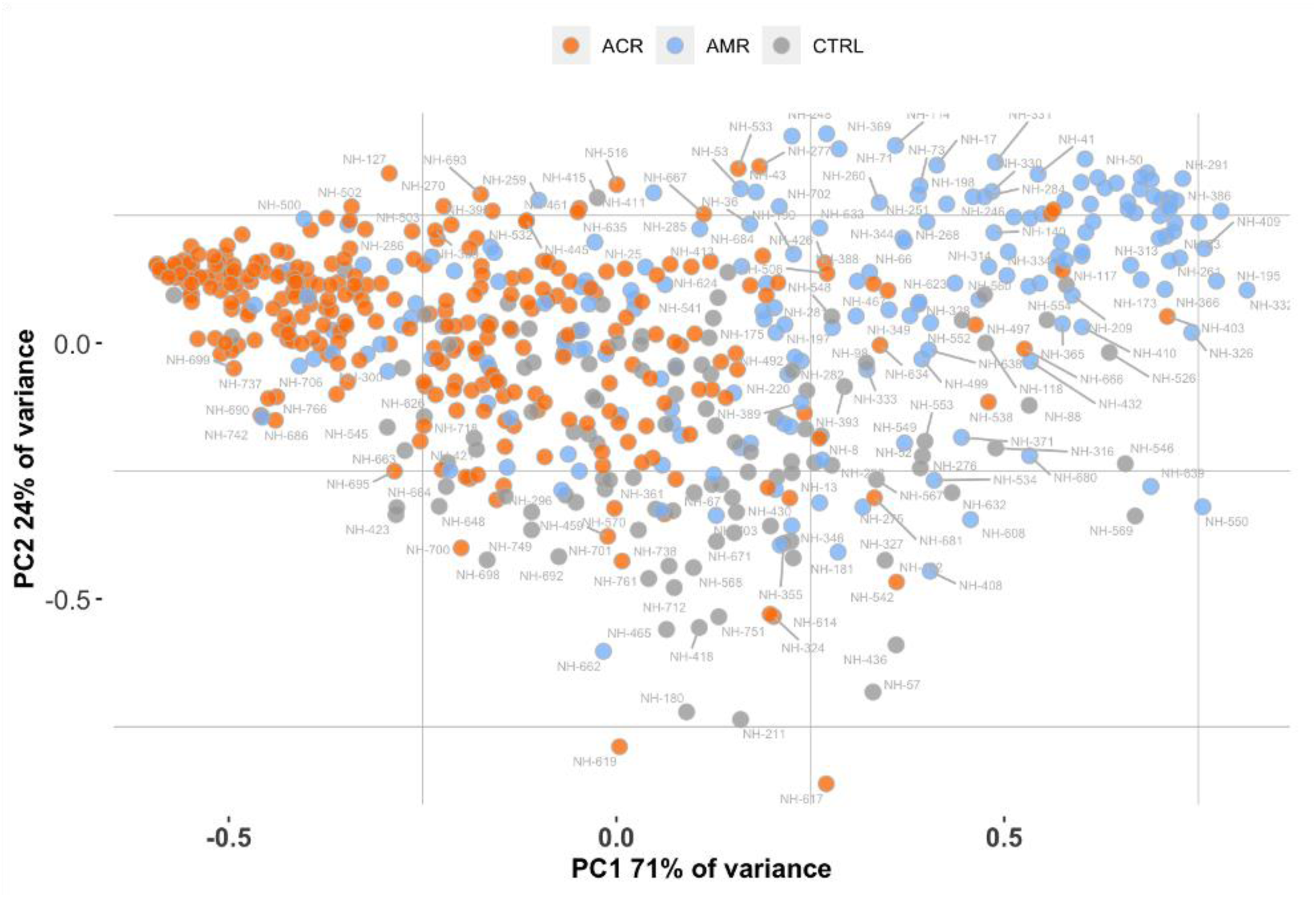
Principal component analysis of heart allograft biopsies. The principal component analysis was derived from molecular scores. Each dot represents one endomyocardial biopsy sequenced passing the quality criteria. Molecular AMR clusters are localized in the upper right of the graph, while ACR attributed biopsies for the molecular scores are localized in the upper left. Biopsies are projected in the PCA space in accordance with each molecular classifier. Histopathological labeling defines AMR (orange dots) ACR (light blue dots) and non-rejection (dots). The axes showed the correlation of individual molecular scores according to the PC1 and PC2, having a great variance of the 72% explained. ACR, acute cellular rejection; AMR, antibody-mediated rejection; PC, principal component.

Overall, the statistical performance of the molecular model to detect AMR in the training set showed a ROC-AUC = 0.832, PR-AUC = 0.763, with a Brier score = 0.141 (**Table 3**, **Figure 5A**). The performance of the molecular model to detect ACR showed comparable metrics: ROC-AUC = 0.849, PR-AUC = 0.849, Brier score of 0.159, F1 score = 0.76 (**Table 3**, **Figure 5C**).

**Figure 5.**
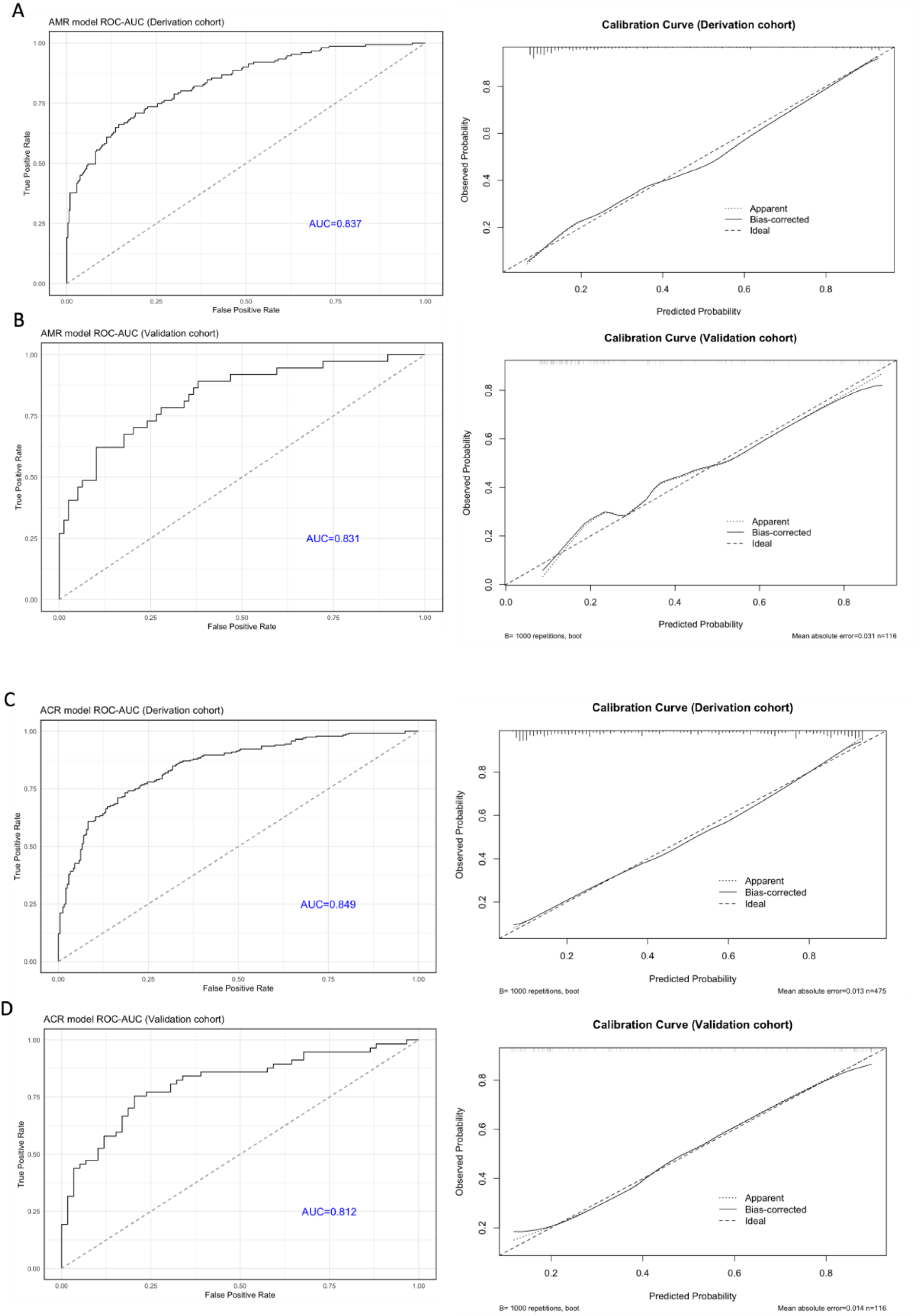
**Evaluation of model performance evaluation: discrimination and calibration curves of molecular models for detecting antibody-mediated and acute cellular rejection**. **Panel A and B**: AMR model performance in the derivation and validation sets, respectively. **Panel C and D**: ACR model performance in the derivation and validation sets, respectively. **Left-hand side**: ROC-AUC curves illustrate the performance assessment of molecular-based models. Discrimination of the AMR and ACR model was excellent, both in the derivation and the validation sets (all AUC > 0.80). **Right-hand side**: Calibration curves. Calibration was adequate for AMR and ACR models in both sets reflecting the reliability of probabilistic predictions made by the molecular diagnostic models. AUC, area under the curve; ROC, receiver operating characteristic.

**Table 3.** Molecular diagnostic model metrics for rejection profiles in heart transplantation in training and validation set. AMR and ACR model performance metrics are assessed for both training and validation set. The precision recall represents the proportion o f positive biopsies in each cohort.

|  | AMR |  |  |  |  |  |  |  | ACR |  |  |  |  |  |  |  |
| --- | --- | --- | --- | --- | --- | --- | --- | --- | --- | --- | --- | --- | --- | --- | --- | --- |
|  | LR top 20 |  | LR MRMR |  | LR MRMR CV |  | RF |  | LR top 20 |  | LR MRMR |  | LR MRMR CV |  | RF |  |
|  | Training | Validation | Training | Validation | Training | Validation | Training | Validation | Training | Validation | Training | Validation | Training | Validation | Training | Validation |
| **AUC** | 0.837 | 0.831 | 0.819 | 0.797 | 0.818 | 0.785 | 0.790 | 0.781 | 0.849 | 0.812 | 0.833 | 0.812 | 0.833 | 0.812 | 0.823 | 0.802 |
| **CI low** | 0.798 | 0.75 | 0.778 | 0.706 | 0.776 | 0.691 | 0.744 | 0.690 | 0.814 | 0.732 | 0.797 | 0.732 | 0.796 | 0.731 | 0.797 | 0.732 |
| **CI high** | 0.876 | 0.913 | 0.861 | 0.888 | 0.860 | 0.880 | 0.836 | 0.873 | 0.883 | 0.892 | 0.870 | 0.893 | 0.871 | 0.892 | 0.870 | 0.893 |
| **PRAUC** | 0.761 | 0.749 | 0.745 | 0.713 | 0.747 | 0.703 | 0.721 | 0.665 | 0.849 | 0.827 | 0.829 | 0.822 | 0.829 | 0.822 | 0.829 | 0.822 |
| **Accuracy** | 77.684 | 81.034 | 75.368 | 79.310 | 76.211 | 74.138 | 77.053 | 73.276 | 77.474 | 77.586 | 77.895 | 76.724 | 77.821 | 76.654 | 77.894 | 76.723 |
| **PPV** | 63.314 | 74.194 | 58.586 | 69.697 | 60.556 | 57.447 | 64.384 | 55.769 | 78.539 | 78.182 | 77.974 | 72.727 | 77.974 | 72.727 | 77.977 | 72.723 |
| **TPR** | 70.861 | 62.162 | 76.821 | 62.162 | 72.185 | 72.973 | 62.252 | 78.378 | 74.138 | 75.439 | 76.293 | 84.211 | 76.295 | 84.211 | 76.281 | 84.211 |
| **TNR** | 80.864 | 89.873 | 74.691 | 87.342 | 78.086 | 74.684 | 83.951 | 70.886 | 80.658 | 79.661 | 79.424 | 69.492 | 79.424 | 69.491 | 79.423 | 69.493 |
| **FDR** | 36.686 | 25.806 | 41.414 | 30.303 | 39.444 | 42.553 | 35.616 | 44.231 | 21.461 | 21.818 | 22.026 | 27.273 | 22.025 | 27.272 | 22.026 | 27.271 |
| **FPR** | 19.136 | 10.127 | 25.309 | 12.658 | 21.914 | 25.316 | 16.049 | 29.114 | 19.342 | 20.339 | 20.576 | 30.508 | 20.576 | 30.508 | 20.578 | 30.509 |
| **F1** | 66.875 | 67.647 | 66.476 | 65.714 | 65.861 | 64.286 | 63.300 | 65.169 | 76.275 | 76.786 | 77.124 | 78.049 | 77.122 | 78.049 | 77.120 | 78.044 |
| **Brier score** | 0.142 | 0.146 | 0.147 | 0.158 | 0.148 | 0.162 | 0.157 | 0.168 | 0.159 | 0.176 | 0.166 | 0.174 | 0.166 | 0.174 | 0.166 | 0.174 |

Abbreviations: LR (logistic regression); mrmr (Maximum Relevance — Minimum Redundancy); RF (Random Forest); CV (cross-validation); AUC (Area Under the ROC Curve);
CI (Confidence Interval); PR-AUC (Precision Recall AUC); PPV (Positive Predictive Value); TPR (True Positive Rate); TNR (True Negative Rate); FPR (False Positive Rate).

### Validation of the gene expression diagnostic system

To assess the generalizability and reproducibility of the gene expression-based rejection system, we tested the AMR and ACR classifier developed in an internal validation set.

Quality controls and normalization were performed independently from the training set and on raw data, AMR and ACR gene expression-based models were applied to the normalized data of the validation set. This validation set included a total of 116 EMB distributed as follows: 37 AMR (pAMR1(I+): n=15; pAMR1(H+): n=12; pAMR2-3: n=10), 57 ACR (1R n=31; 2-3R: n=26), and 22 non-rejection cases.

We found a significant stepwise increase in molecular rejection scores with increasing pathology severity of rejection as assessed by ISHLT working formulations for both ACR and AMR (ACR: p for trend = 1.827E-13, AMR: p for trend = 5.174E-10, **Figure 3B**).

The statistical performance of the molecular model to detect AMR was excellent: diagnostic accuracy = 81.89%, ROC-AUC = 0.844, PR-AUC = 0.742, Brier score = 0.143, and F1 score = 0.70 (**Table 3**, **Figure 5B**). The performance of the molecular model to detect ACR also showed excellent metrics: accuracy = 77.58%, ROC-AUC = 0.812, PR-AUC = 0.827, Brier score of 0.176, F1 score = 0.76 (**Table 3**, **Figure 5D**).

### Learning curves of the models

Learning curves were developed from PR-AUC to illustrate how performance improves over increasing sample size. Models’ learning efficiency is reached around 400 samples size of the cohort (curve’s plateau), showing a convergence of the curves for both training and validation sets (**Supplementary Figure S4**). The statistical performance of molecular models was not limited by the number of samples included in the reference set, thus confirming an appropriately powered study.

### Analysis of discrepancies

Discrepancies between pathology and molecular diagnoses were assessed by comparing observed and predicted classes of rejections. While the overall agreement was excellent, we observed a limited number of discrepancies, particularly related to molecular negative rejection cases (**Figure 6**). Molecular negative ACR cases were more often low-grade rejection (15.28% compared to 12.68%, p<0.05) compared to molecular positive ACR cases. More interestingly, molecular negative AMR cases were less likely to have circulating DSA (8.91% compared to 72.95%, p<0.001) and graft dysfunction (2.57% compared to 15.57%, p<0.005), and more likely to be low-grade rejection (17.21% compared to 15.92%, p<0.05) than molecular positive AMR cases. On the other hand, control biopsies classified as molecular AMR had more frequently circulating DSA (14.45% compared to 8.69, p<0.05) and graft dysfunction (3.47% compared to 1.27 %, p<0.05) than molecular negative control biopsies. Taken together, these results suggest that the molecular analysis may capture sub-histological critical information defining subgroups of molecular-negative rejection cases at lower risk and molecular-positive control cases at higher risk.

**Figure 6.**
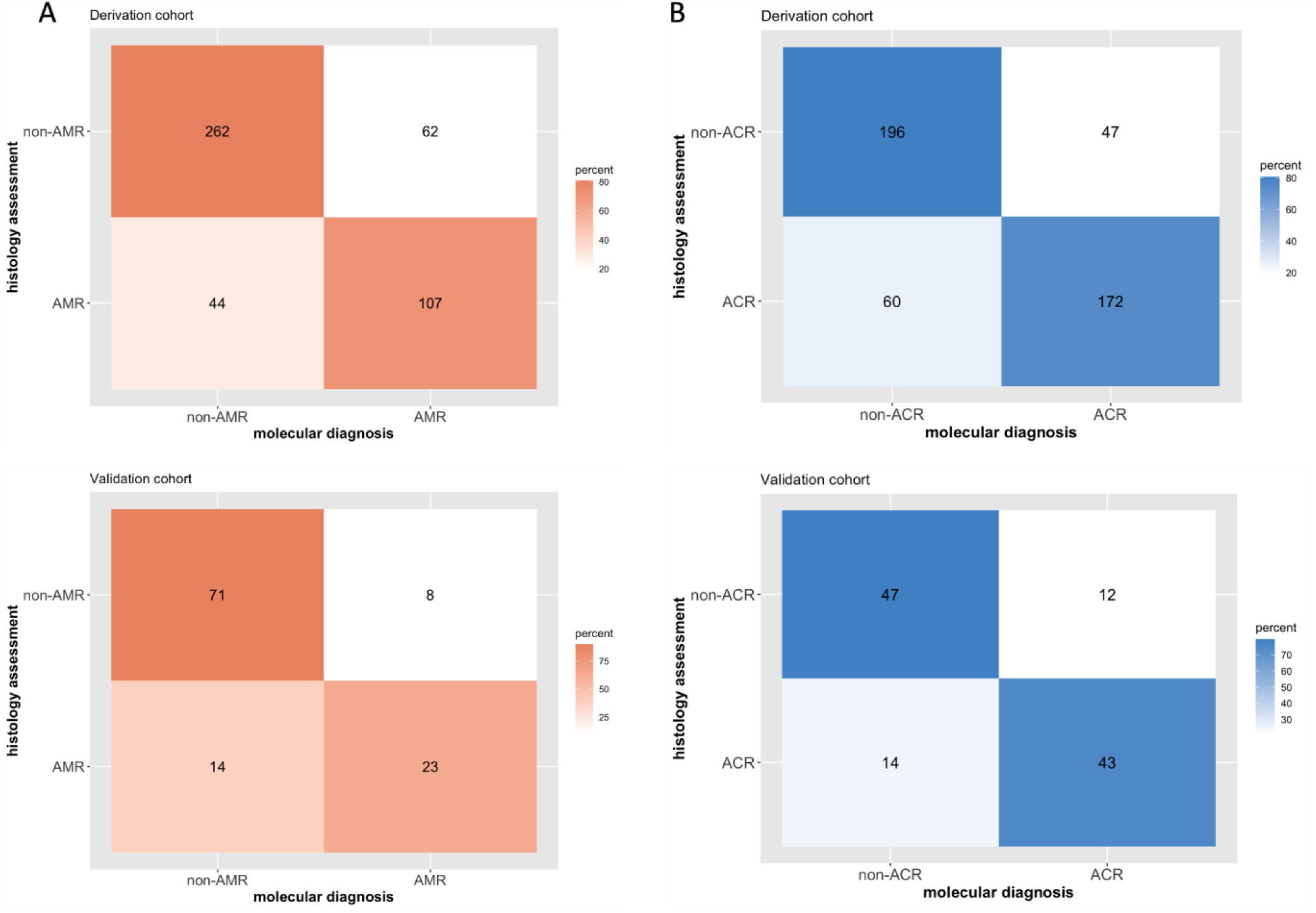
Confusion matrices for antibody-mediated and acute cellular rejection molecular diagnostic models. Discrepancy analysis evaluates the correct attribution of the diagnostic category in the derivation and validation set and identifies discrepant cases for case reconciliation. The confusion matrix shows the actual class labels on the vertical axis (histology assessment) and the predicted class labels on the horizontal axis (molecular diagnosis) for the AMR model (**Panel A**) and ACR model (**Panel B**). Each cell in the matrix quantifies the count of patients’ biopsies that were classified accordingly. Specifically, the diagonal elements (dark colors) represent the correctly predicted instances for each class, while off-diagonal elements (light colors) represent discrepant cases. ACR, acute cellular rejection; AMR, antibody-mediated rejection.

### Automated reporting of gene expression-based classifier for AMR and ACR

To provide clinicians and pathologists with a clinical companion tool, we developed a dedicated molecular report that computes quality control assessment, normalizes and generates predictive molecular scores for any new biopsy (**Supplementary** Figure 5). The report includes all molecular rejection probability scores (AMR, ACR, no rejection), together with the spatial contextualization of the new EMB among the molecular clusters of rejection from the development set (principal component analysis), as well as the main pathophysiological pathways involved, and the k-nearest neighbors (KNN) related to the closest 20 pathology diagnosis. The report includes probabilistic scores instead of binary cut-offs to improve the interpretability of the molecular scores. A range of estimates of the probability derived from the IQR of each score is provided (low, medium, high-risk).

### Sensitivity analyses

Different sensitivity analyses were performed to demonstrate the robustness of our findings across various clinical scenarios and subpopulations.

In our study, 79 EMB (13.5%) were for-cause biopsies compared to 506 (86.5%) protocol EMB and 355 EMB (60.1%) were conducted within the first year post-transplant. Statistical performances of both AMR and ACR models remained adequate and stable regardless of the clinical indication of EMB and time post-transplant. Calibration curves are provided in **Supplementary Figure S6**.

Gene selection methods, including Maximum Relevance - Minimum Redundancy feature selection and random forest-derived models for AMR and ACR, did not show noteworthy improvement in diagnostic performance, despite consistently reducing the number of input variables compared to the linear model (**Table 3**).

In our cohort approximately 30% of patients had multiple biopsies, with a maximum of three EMB analyzed per patient. To account for the potential effect of repeated biopsies from the same patient, one EMB was randomly selected per patient, and both the top differentially expressed genes and pathways analysis did not highlight noteworthy changes, conserving the gene expression signature detected.

Finally, both the derivation and validation sets showed that rejection class events occurred at a rate of approximately 31.8%, with a moderate imbalance specifically in the AMR rejection class. To investigate and minimize the risk of overlooking this potential minority class, we tested four balancing methods—Upsampling, Downsampling, SMOTE, and ROSE—as part of our sensitivity analyses. All these methods did not significantly improve the prediction performance for AMR models, as the resulting metrics were comparable to those of diagnostic systems developed on the reference training cohort (**Supplementary Table S3**).

## DISCUSSION

In this study, we developed and validated in a highly annotated cohort the first tissue-based molecular diagnostic system dedicated to heart transplantation rejection that combines the use of a consensus gene set and a transcriptome technology applicable to FFPE-EMB. The rejection models of the ACR and AMR molecular classifiers were well correlated with the pathology and the pathology severity of rejection. They achieved good performance for detecting AMR and ACR and correctly identified rejection profiles in both the validation (AMR: PR-AUC=0.742; ACR: PR-AUC=0.827) and the derivation sets (AMR: PR-AUC=0.763; ACR: PR-AUC=0.849). Important assets of this approach (reproducibility, costs, FFPE-based, no need for an extra-core biopsy) make this molecular diagnostic system easily applicable in routine clinical care. The automated molecular report might provide valuable and objective data on the graft status and may serve as a novel companion tool to refine the pathology diagnosis of cardiac allograft rejection.

### Precision diagnosis: an unmet medical need in the field of cardiac allograft rejection

The pathologic evaluation of an EMB remains for decades the unique approach to diagnose cardiac allograft rejection. Besides, in heart transplantation, international working formulations do not consider any clinical (graft function) or biological (circulating anti-HLA DSA) parameters for the diagnosis of rejection making the accuracy of the pathology assessment even more crucial. However, major barriers of this exclusively pathology-based strategy limit our ability to provide a precise and reliable diagnosis of rejection, including the use of semiquantitative scales that oversimplify complex phenotypes. While the number of EMB is decreasing with the clinical application of non-invasive biomarkers of rejection (PMID: 33435695, 24944192), there is a need for a deeper tissue-based analysis to improve the diagnostic yield from endomyocardial biopsies (PMID: 32476272). In kidney transplantation, whole-transcriptome studies have defined the molecular phenotypes of allograft rejection and parenchymal injury and contributed to better understanding the pathophysiology underlying rejection processes (PMID: 33107191). The application to heart transplantation of kidney-rejection molecular classifiers represented the first attempt to apply gene expression profiling to refine the diagnosis of cardiac rejection (PMID: 28148598, PMID: 28662985).

As soon as 2013, the revision of the Banff kidney classification for diagnosing AMR added molecular assessment of transcripts indicative of endothelial injury in the renal allograft biopsy as a potential diagnostic criterion, reflecting the need for a more precise evaluation of the graft status (PMID: 24472190). At that time, the lack of agreement on the transcripts to be measured and how to measure them limited the clinical application but further data and revisions contributed to refine the place of the molecular diagnosis (PMID: 27862883). Particularly, the Banff Molecular Diagnostics Working Group, using a data-driven process, designed a consensus 770-gene set (Banff Human Organ Transplantation - B-HOT, PMID: 32428337) that includes the most pertinent genes related to rejection, tolerance, viral infections, and innate and adaptive immune responses discovered and validated in kidney, liver, heart, and lung transplant biopsies.

### Clinical relevance of molecular approaches applicable to FFPE biopsies

Important drawbacks have limited the routine application in daily transplant care of fresh or frozen tissue-based molecular approaches. Particularly, the need for an extra-core sample dedicated to the molecular analysis increases the risk of procedure-related complications and sampling bias and does not allow the oversight of the pathology to exclude the influence of nonspecific lesions such as fibrosis, fibrin clot, previous biopsy site, pericardial tissue, or Quilty effect (PMID: 32476272). Additionally, the limitations of the whole-transcriptome approach related to low reproducibility, variation due to cDNA conversion, amplification steps, transcript labeling, and probe redundancy consistently hampers the integration of such approach in routine transplant care. For these reasons, the Banff heart group recommended the use of FFPE-based technologies to evaluate the clinical applicability of molecular diagnostic in heart transplantation (PMID: 32476272, 37838218).

Interestingly, the B-HOT panel is a commercially available panel that uses the NanoString platform, which can provide an absolute quantification of transcripts from FFPE samples, allowing the analysis of all available biopsy fragments, thus minimizing sampling bias issues. The nCounter system (NanoString) is highly adaptable, reproducible, and amenable to collaborative efforts between centers (PMID: 32476272). The platform is well-adapted to intermediate numbers of targets, does not require cDNA conversion, and raw data files are 200 times smaller than for microarray, which represent major advantages to improve diagnostic accuracy while reducing complexity, cost, and turn-around time, necessary for the implementation of transcriptome sequencing into the routine rejection diagnosis pipeline (PMID: 37745639).

Our group recently showed *in silico* that the B-HOT panel-based gene expression analysis was able to capture biologically and clinically relevant genes, pathways and networks reflecting the key pathophysiological mechanisms of AMR in heart transplantation. A proof-of-concept study that supported the clinical use of the B-HOT panel as a reliable proxy to whole-transcriptome analysis for gene expression profiling of heart allograft biopsies (PMID: 37745639). The present study represents the next step forward by demonstrating the reliability of targeted molecular diagnostic classifiers that may overcome whole transcriptome limitations. Compared to previous molecular diagnosis systems, our approach presents several assets, including (i) a robust study design dedicated to HTx that relies on an international multicenter approach with a core lab for central review of biopsies and a large sample size of biopsies reflecting a wide spectrum of transplant pathology, (ii) significant technical improvements with the use of a consensus gene set and a transcriptome technology applicable to FFPE-EMB allowing molecular analysis on the same FFPE tissue used for histology assessment, (iii) a robust statistical approach with the development and validation of molecular classifiers strongly associated with the severity of rejection as assessed by the pathologist, which performance was not limited by sample size, providing an objective molecular diagnosis of rejection, and (iv) the direct comparison of histology and molecular diagnoses through the use of confusion matrices and the analysis of diagnostic discrepancies. These important aspects may contribute to minimizing discrepancies between histology and molecular diagnosis and reduce overdiagnosing AMR, two important limitations observed with previous molecular approaches (PMID: 38294835).

Improved diagnostic precision plays a critical role in differentiating between alloimmune-mediated injury and non-specific injury, thereby optimizing the equilibrium between over- and under- immunosuppression. This is particularly crucial in early rejection stages, before it is visible to histology assessment or in cases with minimal molecular rejection signs following the adapted treatment. Discordances between current gold standard histological classification and molecular profiling often reveal shortcomings in both methods, highlighting the necessity and the added value of a comprehensive diagnostic approach. This multimodal diagnostic approach could significantly enhance the accuracy of rejection diagnoses and ultimately improve patient outcomes by optimizing immunosuppressive therapy and reducing the risk of over- or under-treatment.

### Limitations and future developments

Our study is the first endeavor to develop and validate the performance of B-HOT gene panel based predictive models for cardiac allograft rejection. The introduction of an automated reporting system can streamline the assessment of the clinical utility of a gene-expression based decision support tool in transplant patient care. However, its potential long-term impact on graft survival requires the development of multicenter prospective studies. Discordant cases highlight the limit of both gene expression based and histological diagnostic methods. As the true disease state is unknown and graft loss and treatment response are unreliable outcomes, assessing the added value of molecular diagnostics in cardiac transplantation is challenging. The successful integration of gene expression analysis into clinical practice requires the adoption of standardized protocols, reproducible technologies and careful result interpretation alongside all patient related, immunological, histological, and diagnostic information. By thoroughly considering all the relevant clinical parameters in multivariate gene expression models, precision diagnostic in cardiac transplant care can be further optimized.

## Conclusion

Using targeted FFPE-based transcriptome technology with a gene panel designed to detect pivotal pathophysiological pathways in transplant rejection, we developed and validated a molecular diagnostic system with improved reproducibility and reliability, specifically designed for cardiac allograft rejection. This approach is easily applicable in clinical practice as a pathology companion tool and has the potential to refine the diagnosis of rejection.

### Data Availability Statement

The data that support the findings of this study are available from the corresponding author upon reasonable request.

## Data Availability

All data produced in the present study are available upon reasonable request to the authors

## Abbreviations

ACR: Acute Cellular rejection
AMR: Antibody-mediated rejection
AUC: Area Under the ROC Curve
B-HOT: Banff Human Organ Transplant Panel
DEG: Differentially Expressed Genes
DSA: Donor-Specific Antibodies
EMB: Endomyocardial Biopsies
FDR: False Discovery Rate
FFPE: Formalin Fixed Paraffin Embedded
ISHLT: International Society of Heart and Lung Transplantation
PCA: Principal Component Analysis
PR-AUC: Precision Recall-AUC
ROC: Receiver Operating Characteristic

## SUPPLEMENTARY MATERIAL AND METHODS

### Supplementary Methods

#### 1. Cohort selection

Due to the low prevalence of rejection in contemporary cohorts, a selection of cases and controls was mandatory to avoid the analysis of numerous non-rejection cases, which represent more than 90% of biopsies in our cohorts (PMID: 36200456). However, a strict study design has been applied to mitigate the risk of selection bias. Allograft biopsies were selected at each transplant center according to the following common criteria:

#1- First, we pre-selected biopsies (i) from adult heart transplant recipients (>18 years old), (ii) performed between 1-month (to avoid ischemia reperfusion molecular signals) and 10-year (to avoid chronic allograft injuries molecular signals) post-transplant, (iii) without cardiac allograft vasculopathy grade ≥ 2 on last coronary angiogram and/or prior history of coronary stenting since transplant, (iv) without ongoing cytomegalovirus infection for EMB performed during the first year post-transplant (last viral PCR < 1000 UI/mL).
#2- Second, a predefined number of each type of rejection cases (n≥50) was defined to explore the entire molecular landscape of cardiac allograft rejection (ACR 1R1A, 1R1B-1R2, 2R and 3R; pAMR1(I+), pAMR1(H+), pAMR2-3). To develop clear and stable molecular rejection scores, ambiguous cases including biopsy-negative rejection, mixed rejection, and Quilty cases were excluded. Rejection cases were randomly selected at each transplant center according to the predefined number of each type of rejection required. The experimental design allowed the analysis of more than 1 biopsy per patient.
#3- Third, controls were randomly selected among patients without rejection during follow-up (ACR ≥ 2R, AMR ≥ pAMR1) and with normal graft function at the time of EMB.

#### 2. Histology assessment

Affinity-purified anti-human C4d rabbit polyclonal antibody (Biomedica Gruppe, Vienna) was performed for C4d immunostaining. Sections were retrieved using Citrate buffer solution on microwave (Histos 3; Milestone) and incubated with anti-C4d antibody at a 1/500 dilution at room temperature for 1 hour, with anti-rabbit EnVision (Dako Corporation, Hamburg, Germany) for 30 minutes and, finally, with peroxidase diaminobenzidine (DAB) for 5 minutes. Mayer’s hematoxylin solution was used for 1 minute to counterstain the sections, then dehydrated in alcohol progressively, and mounted on the slide glass with medium mounting (Eukitt; Bioptica). Immunohistochemical (IHC) staining of CD68 was performed routinely and retrospectively on the paraffin sections of each EMBs for monocyte/macrophage evaluation and examined by light microscopy. Monoclonal mouse anti-human CD68, Clone PG-M1 (dilution 1/200; Dako), was retrieved using an ethylene-diamine tetraacetic acid (EDTA) solution on a microwave oven. To discriminate whether CD68 positive macrophages were intra- or extra- capillary additional staining for CD34 antibody (Immunotech) was performed on selected biopsies.

#### 3. Definition of Antibody-Mediated Rejection of Heart Allografts

Histology of EMBs was assessed by 2 expert pathologists. Biopsies were graded according to the most recent recommendation of Society for Heart and Lung Transplantation (PMID: 21555100, PMID: 16297770). Furthermore, immunohistochemistry based on C4d capillary deposition (positive if >50% of the capillaries were labeled) and/or CD68-positive staining (positive if intravascular CD68+ macrophages were present in >10% of the capillaries) were evaluated.

#### 4. RNA isolation

Up to four 20 -μm thick sections were collected from each EMB-FFPE tissue blocks to obtain 100 ng of RNA required for analyses. RNA was isolated and extracted using the tissue RNEasy FFPE kit #73504 (Qiagen, Hilden, Germany). The RNA isolation process can be summarized in three key steps: i) deparaffinization, ii) removal of contaminant (both organic and DNA related); iii) final serial centrifugation to remove salt contaminant and concentrate isolated RNA. The concentration yield and quality of isolated RNA were assessed using NanoDrop 2000 spectrophotometer (Thermo Fisher Scientific Inc., Waltham, MA, USA).

#### 5. Transcriptome analysis

Total RNA from each bioptic sample was hybridised with the nCounter® Human Banff Orga Transplant gene panel (B-HOT, NanoString Technologies, Seattle, WA, USA). This panel was created by the Banff Molecular Diagnostics Working Group, comprising a selection of verified genes from significant, peer-reviewed publications on molecular profiling in transplantation. The consortium-approved B-HOT panel assesses the mRNA expression of 758 target genes, including 12 chosen housekeeping genes for data normalization. This panel covers key pathways and processes linked to damage and rejection in solid organ transplants, as well as innate and adaptive immune responses. A Panel Standard, a pool of artificial DNA oligonucleotides corresponding to the endogenous probes, was included in each run to ensure the internal quality standard for each test. This allowed for the normalization of user, instrument, and lotto-lot variability.

#### 6. Quality control of gene expression sequencing data

Overall, the molecular analysis of 592 (97%) biopsies successfully passed all quality control and normalization steps. We excluded 18 FFPE-EMBs that do not meet quality criteria of quality and purity (1.8-2.00 for A260/30 and 260/280 ratio) as well as Imaging and Binding density, linearity of the positive controls and limit of detection. All the samples passed the QC metrics which were defined as binding density > 0·05 and < 2·25 probes/μm2, % fields of view > 0·75, positive control linearity > 0·95, and sufficient assay efficiency. Per-sample LOD was defined as the mean of the counts of negative control probes for each sample and was ensured by using internal quality control reference probes. Eventual batch effects were evaluated through principal component analysis on overall gene expression data with the highest variance for all the samples included (**Supplementary Material**). Despite having biopsies from different experimental batches and centers, the retrospective reference set showed similar gene expression distribution with no evidence of site-specific batch effects, excluding from subsequent analysis only two biopsies not homogeneously clustered within the overall reference set. The final included biopsies cohort, after exclusion of biopsies for quality control and batch effect comprehend biopsies for: 189 AMR, 289 ACR, 114 non-rejection cases. Elective housekeeping genes (HK) present in the B-HOT gene panel were selected according to average pairwise variation computed with all other B-HOT genes, keeping HK genes with an M value (<0.7) (*i.e.,* ABCF1, G6PD, GUSB, NRDE2, OAZ1).

#### 7. Housekeeping gene assessment

The housekeeping genes are constitutive genes with stable expression in varying conditions, the BHOT gene panel includes 758 endogenous and 12 housekeeping (HK) genes (gene symbols: ABCF1, G6PD, GUSB, NRDE, OAZ1, POLR2A, PPIA, SDHA, STK11IP, TBC1D10B, TBP, UBB). The housekeeping genes function in the gene panel serve two purposes related to: i) QC to remove samples with overall poor quality of sequencing and ii) to assess the technical variation present for normalization procedure. The ideal HK genes showed stable expression with low coefficient of variation, with good correlation with HK genes overall the samples included in the cohort (Nanostring, PMID: 22513995, PMID: 32789507). To select stable housekeeping genes that were used for normalization, *geNorm* algorithm was applied, HK genes were ranked from the most to the least stable, computing the average pairwise variation with all other genes, giving a stability value (M). Genes with the highest M value (>0.7) are then excluded from the normalization pipeline.

#### 8. Normalization for targeted transcriptome data

Gene Expression counting analysis of RNA samples of allograft biopsies was performed using the NanoString nCounter platform. Raw expression data were normalized to corrected eventual technical variation between samples and batches using internal positive and negative reference probes and housekeeping genes using both nSolver and RUV method (Nanostring, PMID: 22513995, PMID: 32789507, PMID: 31114909).

#### 9. Reduced Unwanted Variation

The RUV method is designed to estimate the factors of unwanted variation and adjust for these in the model, considering both the expression values of the samples and those of the negative control transcripts (PMID: 31114909). First upper-quantile normalization was used to scale the distributional differences between lanes. Then, to estimate unwanted technical factors in the sequenced gene expression data obtained, *RUVg* function (RUVSeq package) was applied. *RUVg* approach relies on the identification of a set of negative control genes (genes not expected to be differentially expressed across the samples, *e.g.*, housekeeping genes or spike-ins) not affected by the covariates of interest. RUV methods follow a generalized linear model in which targeted sequenced counts for each endogenous gene are regressed on the covariates of interest, making then a subset of the data to estimate the factors of unwanted variation and adjust the sequencing data. This model was fitted to the counts and the estimated factor of unwanted variation and HK scaling factors.

The original count data were transformed using DESeq2 to stabilize the variance, and the estimated undesirable variation was then subtracted using the removeBatchEffects function of the limma package. Returning then the gene expression set normalized in accordance.

Resulting relative log expression (RLE) plots were implemented to visualize the deviation between raw and normalized counts per gene from the median expression across all samples. Raw counts RLE plot showed unwanted variation in high dimensional gene expression data, subsequently resolved by applying RUV normalization procedure.

#### 10. Feature selection and balancing classes

MRMR and RF feature selections were developed for AMR models, despite consistently reducing the input feature in the linear model, do not show noteworthy improvement in the diagnostic performance. Thus, consistently presenting PR-AUC of 0.71 for MRMR feature selection, with MRMR CV PR-AUC = 0.70 (Table 2-3). It is possible to notice comparable results in the feature selection adjustment models for ACR. MRMR and RF showed similar PR-AUC with a median of 0.821 and a calibration Brier score of 0.173. We showed similar behavior for ACR models combined with feature selection methods.

#### 11. Correlation and multicollinearity

Gene correlation was investigated to rank the dependency of differentially expressed genes according to Pearson score and visualized using heatmaps (PMID: 21576180). We investigated potential multicollinearity conundrum computing the variance inflation factor for each gene included in each rejection signature (**Supplementary Methods**).

Heatmaps associated with gene correlation were performed to visualize and rank the dependency of the feature selected within each model computed. op ranked genes for AMR showed absence of dependency and correlation within the variable. While for ACR we have a higher correlation between the set of features selected. Variable inflation factors (VIF) were computed to assess potential gene correlation between the features selected, showing for both AMR and ACR genes selected a VIF below the recommended index of 5. Finally, MRMR and RF approaches were implemented as additional feature selection methods, resulting in reducing consistently the set of selected genes. Having 7 genes for AMR-associated model and 5 genes ACR-associated, optimizing the correlation coefficient and visualization through heatmap.

#### 12. Class imbalance

To minimize the risk of overlooking the minority class, four balancing methods were applied (upsampling, downsampling, SMOTE and ROSE) on the reference set trying to balance the minority class. The most common approach to deal with imbalance dataset involves altering the class distribution to obtain a balanced dataset. To address this concern, the reference dataset was balanced by resampling the rejection classes using at first two canonical approaches: the Downsampling or Undersampling method, referring to the reducing of the majority class samples to balance with the minority class label (function upSample); or the Upsampling or Oversampling procedure, a methods that create artificial or duplicate data of the minority class to balance the label attributed (function downSample). Furthermore, other two methods for balancing minority class are applied to the reference set, the first relying on the Synthetic Minority Oversampling Technique (SMOTE) in which for each rare training observation, new examples are generated by randomly choosing points that lie on the line connecting the rare observation to one of its nearest neighbors in the feature space; and the Random Over Sampling Examples (ROSE) based on the generation of new artificial data from the classes, according to a smoothed bootstrap approach.

**Supplementary Figure S1.**
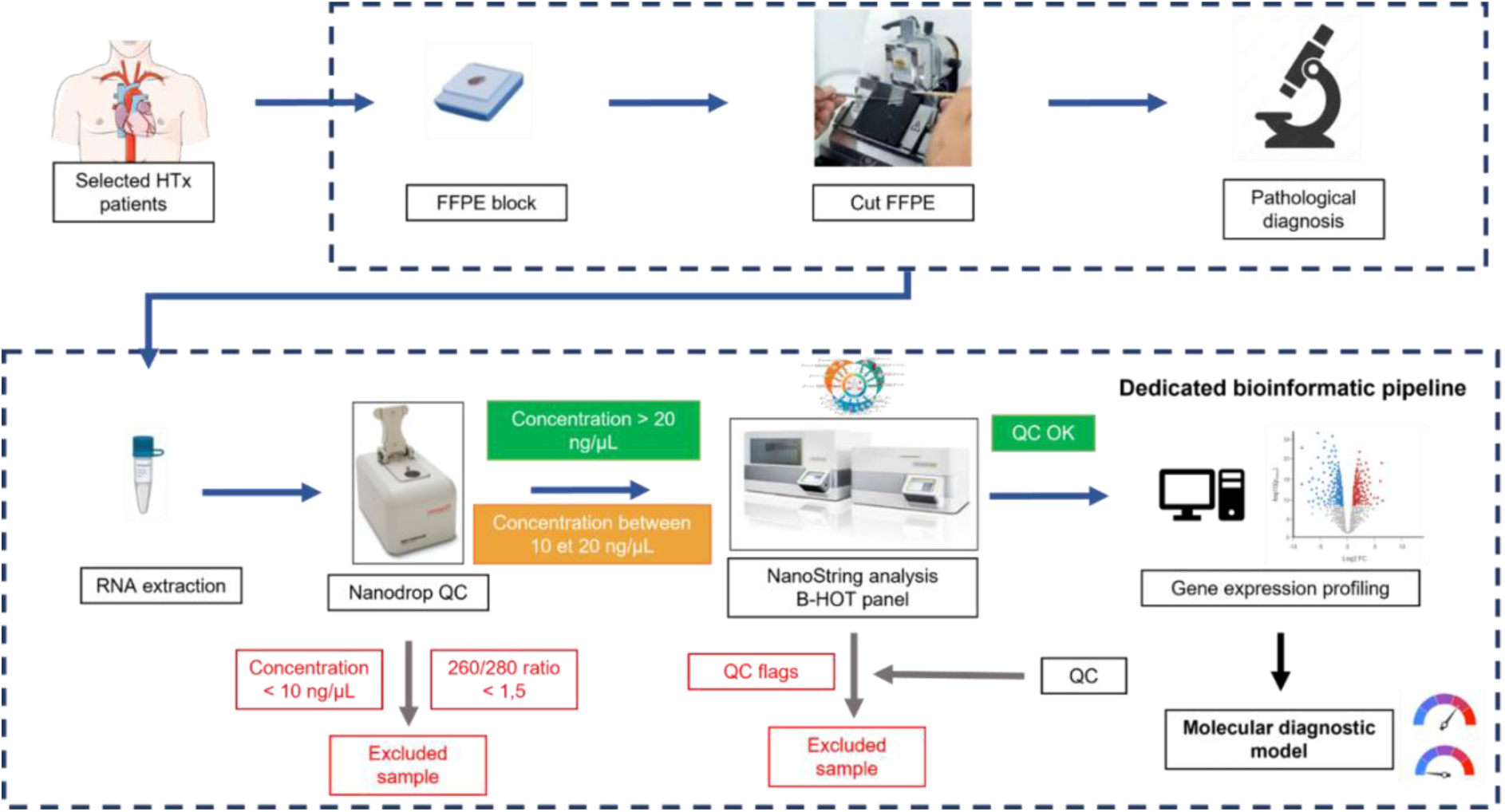
Experimental workflow for molecular assessment.

**Supplementary Figure S2.**
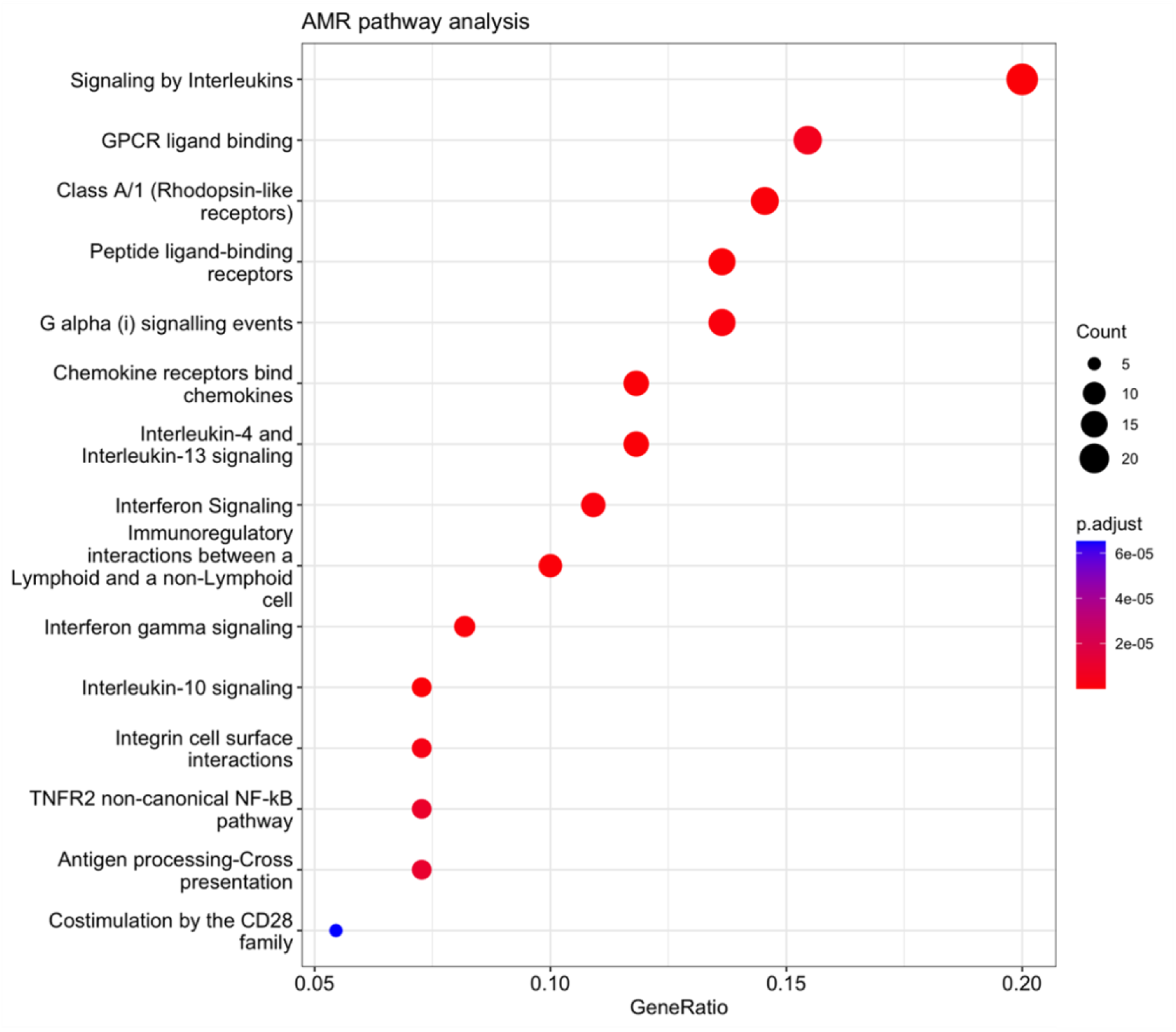
Top ranked pathways associated with AMR. Dot plots show the top 15 enriched pathways based on significant differentially expressed genes (FDR<0.05) associated with AMR. The top 15 significant pathways are plotted in order of gene ratio (number of genes associated with the given pathway divided by the total number of genes analyzed). The size of the dots represents the number of genes in the significantly differentially expressed gene list associated with the pathway and the color intensity represents the p-value.adjusted for false discovery rate.

**Supplementary Figure S3.**
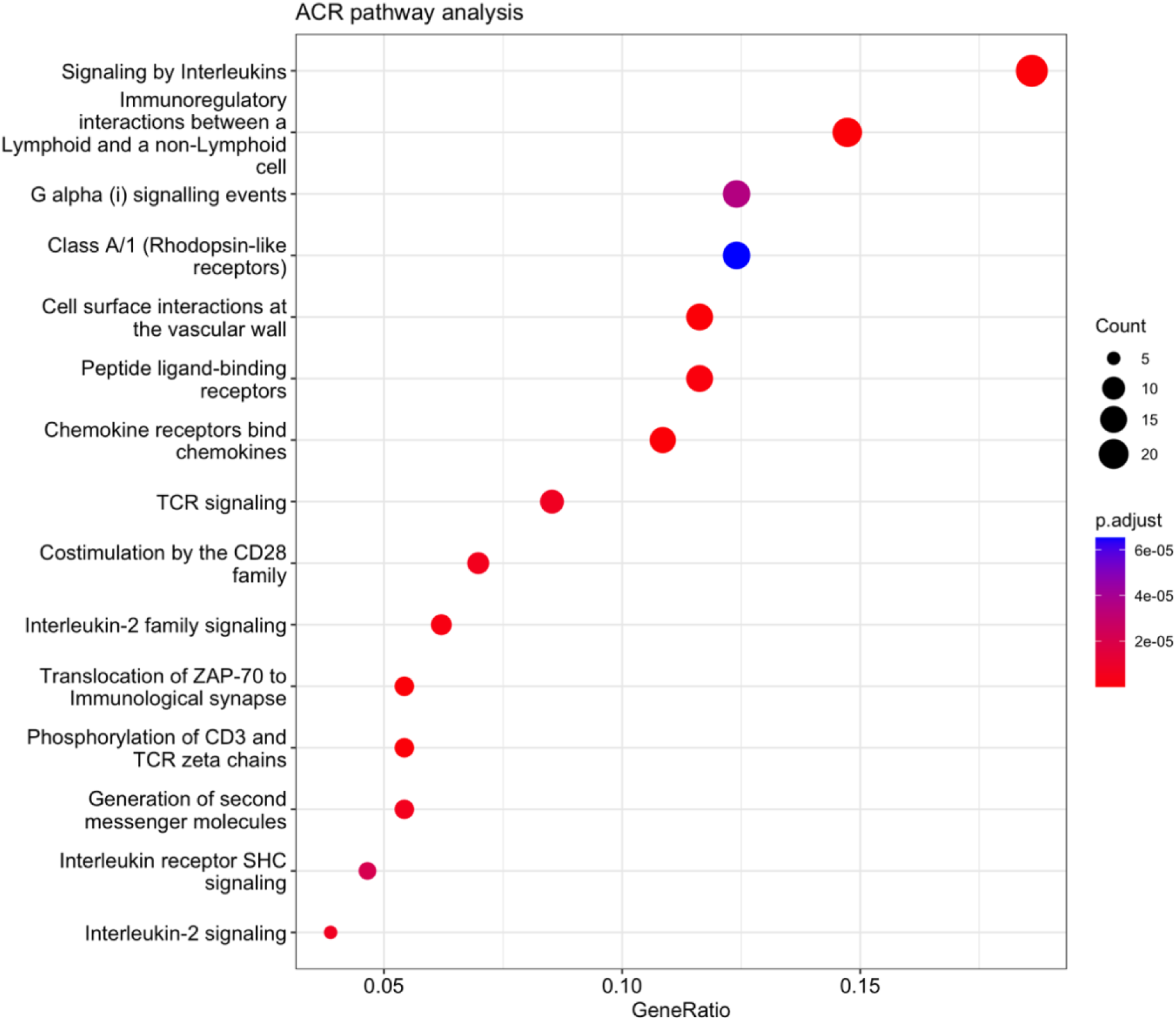
Top ranked pathways associated with ACR. Dot plots show the top 15 enriched pathways based on significant differentially expressed genes (FDR<0.05) associated with ACR. The top 15 significant pathways are plotted in order of gene ratio (number of genes associated with the given pathway divided by the total number of genes analyzed). The size of the dots represents the number of genes in the significantly differentially expressed gene list associated with the pathway and the color intensity represents the p-value.adjusted for false discovery rate.

**Supplementary Figure S4.**
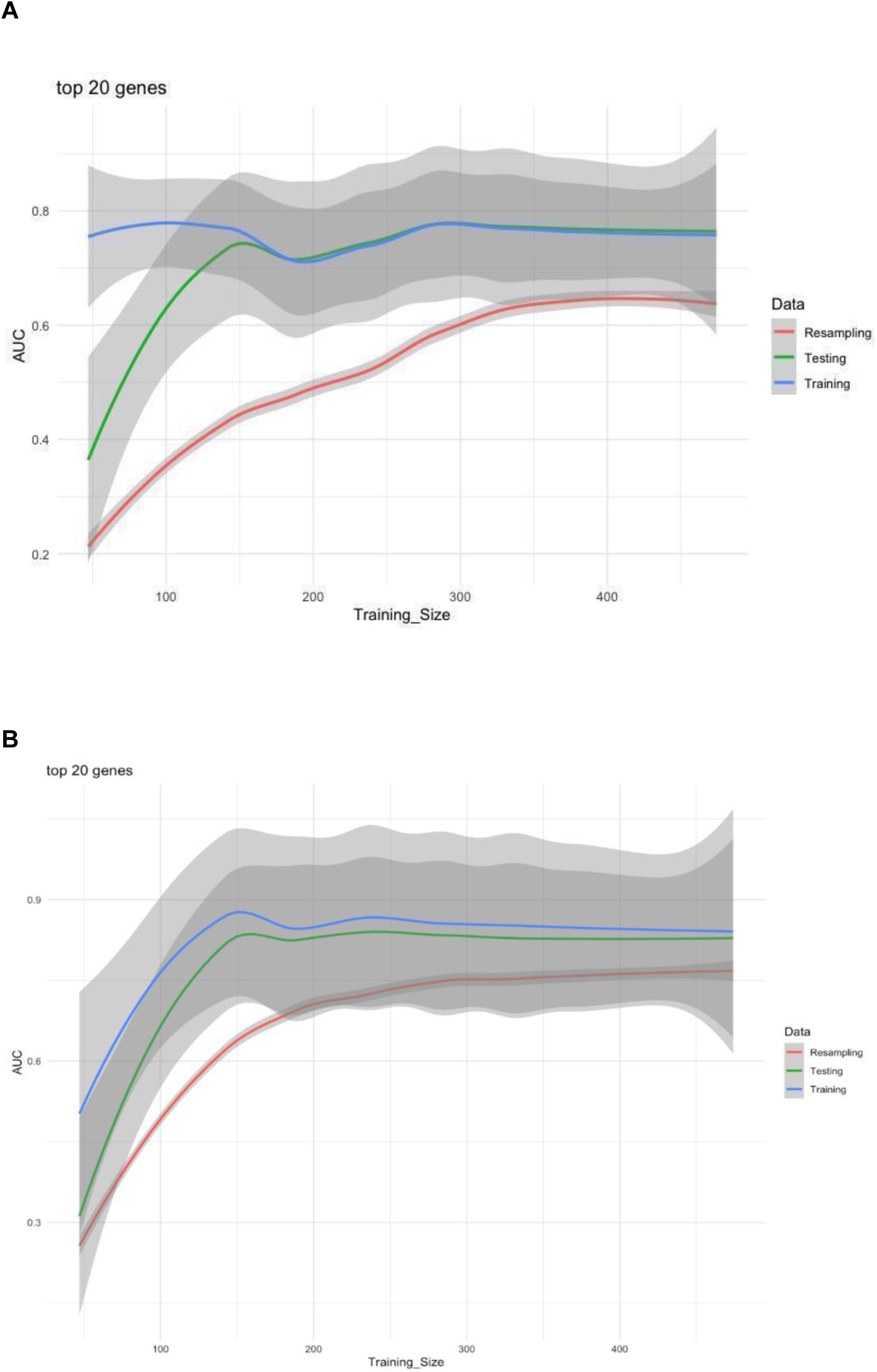
Learning curve of AMR and ACR BHOT-molecular based model. The learning curve graph demonstrates the model’s performance evolution across training and validation epochs for AMR (Panel A) and ACR (Panel B) models respectively. The x-axis represents epochs or iterations, while the y-axis depicts a chosen performance metric to assess accuracy or loss, in our case PR-AUC. Learning curves allow for the assessment of convergence, overfitting, or underfitting. It aids in determining optimal training epochs, identifying model behavior, and guiding adjustments to enhance predictive performance. In our study we can demonstrate that both the size of the training and test set are consistent and adequate to develop and assess the B-HOT base molecular diagnostic model.

**Supplementary Figure S5.**
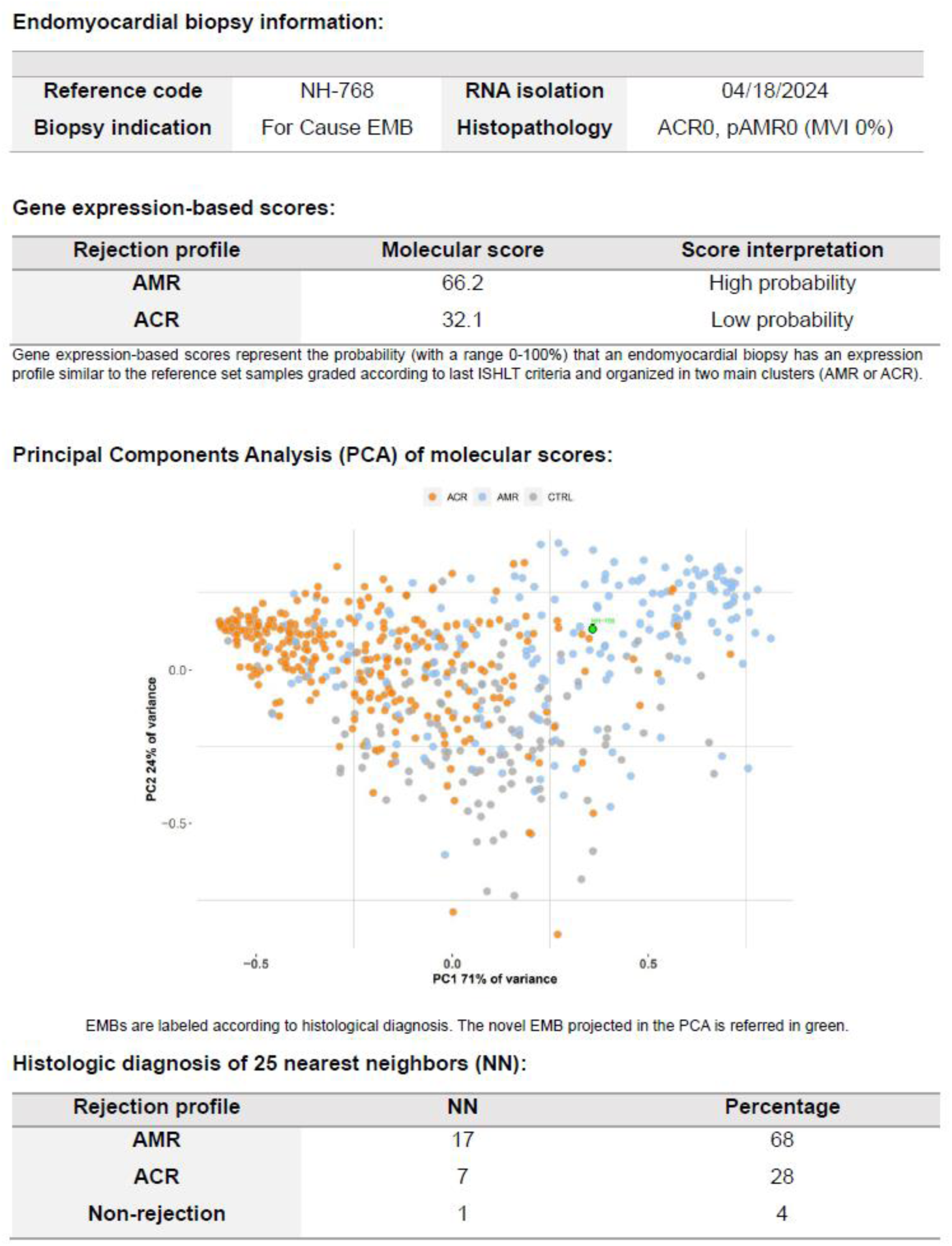
Molecular report for heart transplant rejection. Representative screenshot of the gene expression-based report derived from the molecular classifiers, demonstrating the potential clinical utility of targeted transcriptome approach from the present multicenter study. Histology-based diagnosis is reported for a challenging case, together with AMR, ACR scores, interpretation, PCA e nearest neighbor analysis.

**Supplementary Figure S6.**
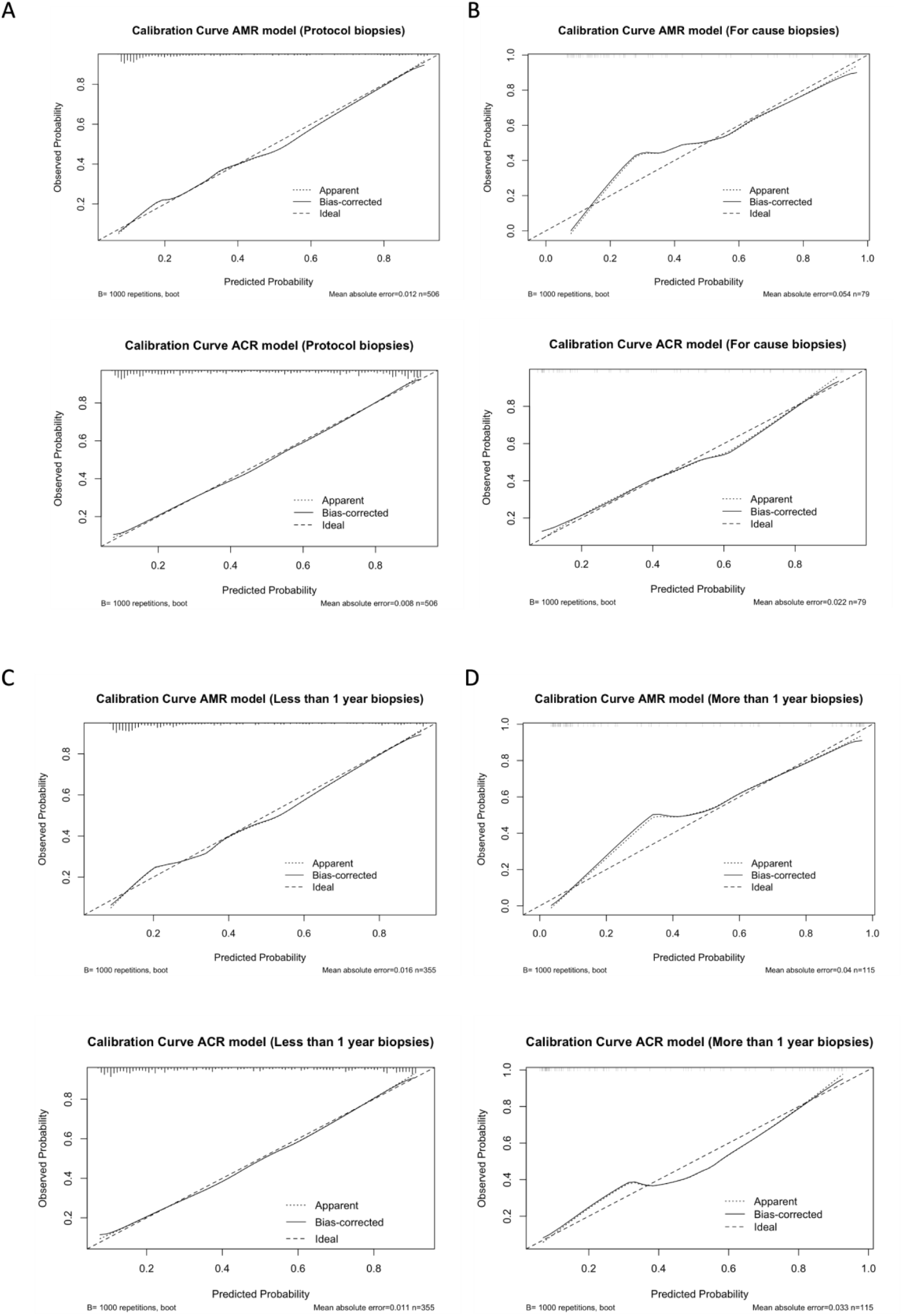
Sensitivity analysis for AMR and ACR models. Calibration curves were adequate for AMR and ACR models accounting for biopsy type protocol (Panel A) and for cause (Panel B), as well as for biopsy performed within the 1 -year (Panel C) or after (Panel D), demonstrating the reliability of the molecular diagnostic models.

**Supplementary Table S1.** Top 20 significant genes associated with antibody-mediated rejection in heart allografts.

| Gene | Gene name | log2FC | p-value | FDR p-value |
| --- | --- | --- | --- | --- |
| PLA1A | phospholipase A1 member A | 0.96417 | 2.89E-36 | 2.10E-33 |
| CXCL11 | C-X-C motif chemokine ligand 11 | 1.09031 | 1.83E-27 | 1.33E-24 |
| PECAM1 | platelet and endothelial cell adhesion molecule 1 | 0.40679 | 1.48E-25 | 1.08E-22 |
| CCL19 | C-C motif chemokine ligand 19 | -1.00396 | 1.93E-19 | 1.40E-16 |
| PLAT | plasminogen activator, tissue type | 0.47318 | 1.19E-18 | 8.64E-16 |
| TM4SF18 | transmembrane 4 L six family member 18 | 0.42004 | 5.62E-18 | 4.09E-15 |
| PHEX | phosphatase regulating endopeptidase homolog X-linked | -0.76565 | 6.29E-17 | 4.58E-14 |
| LILRB1 | leukocyte immunoglobulin like receptor B1 | 0.42743 | 2.83E-16 | 2.06E-13 |
| FCGR3AIB | Fc fragment of IgG receptor IIIb | 0.62000 | 4.14E-16 | 3.01E-13 |
| CX3CL1 | C-X3-C motif chemokine ligand 1 | 0.48819 | 5.11E-16 | 3.72E-13 |
| NOS3 | nitric oxide synthase 3 | 0.27418 | 1.10E-14 | 7.99E-12 |
| CLEC4C | C-type lectin domain family 4 member C | -0.58463 | 1.47E-14 | 1.07E-11 |
| LTB | lymphotoxin beta | -0.60692 | 5.47E-14 | 3.98E-11 |
| IL1R2 | interleukin 1 receptor type 2 | 0.71641 | 1.45E-13 | 1.06E-10 |
| ROBO4 | roundabout guidance receptor 4 | 0.33836 | 4.62E-13 | 3.37E-10 |
| HYAL2 | hyaluronidase 2 | 0.30476 | 4.77E-13 | 3.47E-10 |
| BATF | basic leucine zipper ATF-like transcription factor | -0.34916 | 7.95E-13 | 5.79E-10 |
| GATA3 | GATA binding protein 3 | -0.29516 | 1.02E-12 | 7.43E-10 |
| KLF4 | Kruppel like factor 4 | 0.24884 | 1.80E-12 | 1.31E-09 |
| CAV 1 | caveolin 1 | 0.22343 | 1.55E-11 | 1.13E-08 |

Abbreviations: FDR p-value (p-value adjusted for false discovery rate); FC (fold-change).

**Supplementary Table S2.**
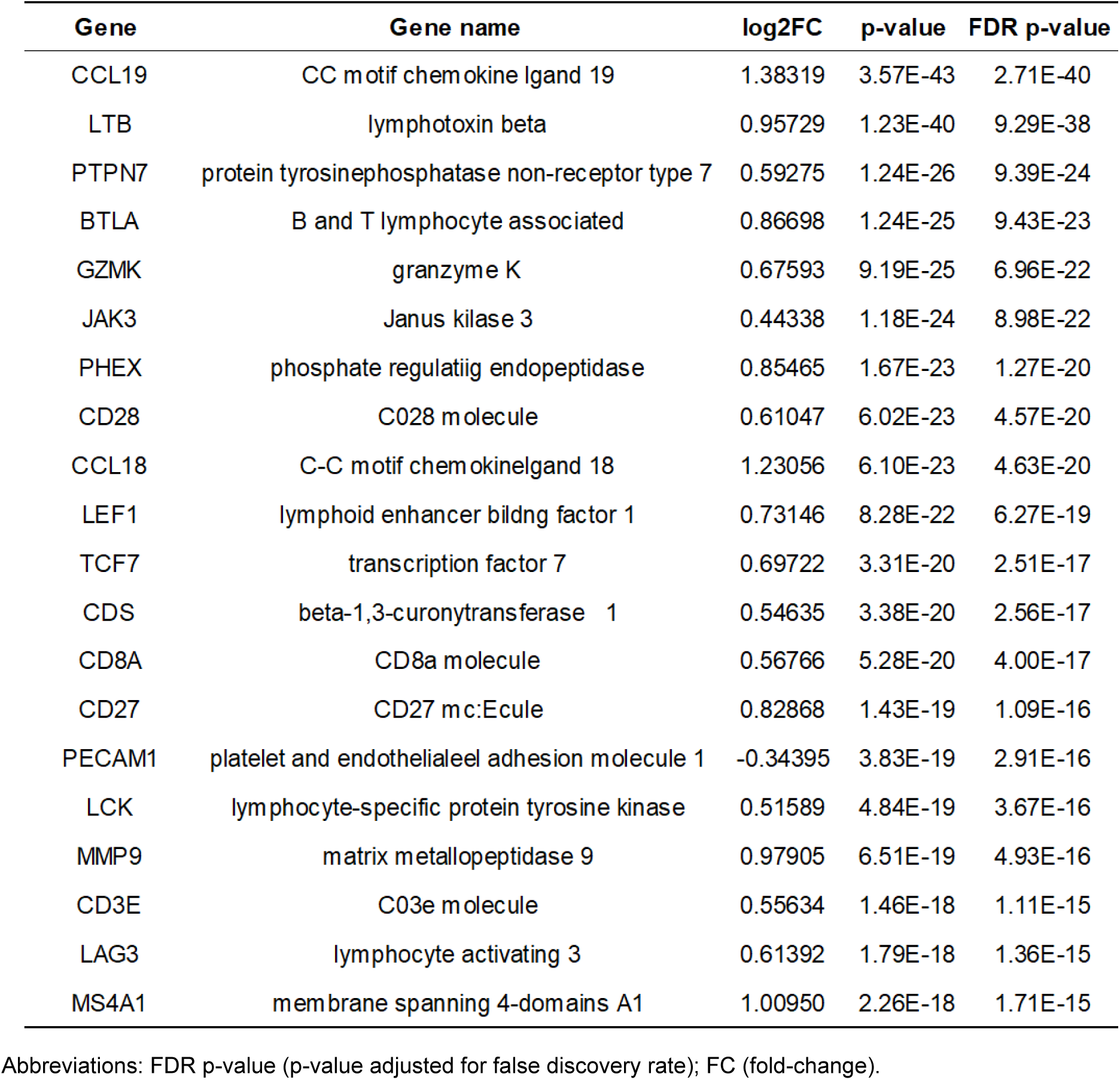
Top 20 significant genes associated with acute cellular rejection in heart allografts.

**Supplementary Table S3.** Balancing minority class sensitivity analysis for molecular diagnostic models for AMR profile in heart transplantation. AMR performance metrics are assessed for both training and validation set applying 4 strategies to balance minority classes (Upsampling, Downsampling, SMOTE, ROSE). The precision recall represents the proportion of positive biopsies in each cohort.

|  | AMR |  |  |  |  |  |  |  |
| --- | --- | --- | --- | --- | --- | --- | --- | --- |
|  | Upsampling |  | Downsampling |  | SMOTE |  | ROSE |  |
|  | Training | Validation | Training | Validation | Training | Validation | Training | Validation |
| **AUC** | 0.833 | 0.848 | 0.832 | 0.837 | 0.830 | 0.830 | 0.824 | 0.823 |
| **CI low** | 0.794 | 0.775 | 0.792 | 0.759 | 0.791 | 0.747 | 0.746 | 0.746 |
| **CI high** | 0.873 | 0.922 | 0.871 | 0.915 | 0.870 | 0.912 | 0.902 | 0.891 |
| **PRAUC** | 0.7566 | 0.7371 | 0.7484 | 0.7306 | 0.7445 | 0.7229 | 0.7081 | 0.7020 |
| **Accuracy** | 78.7368 | 79.3103 | 76.6316 | 81.8966 | 77.0526 | 80.1724 | 75.8621 | 75.8630 |
| **PPV** | 67.1233 | 64.4444 | 61.1111 | 73.5294 | 62.0690 | 66.6667 | 59.1837 | 59.1880 |
| **TPR** | 64.9007 | 78.3784 | 72.8477 | 67.5676 | 71.5232 | 75.6757 | 78.3784 | 78.3788 |
| **TNR** | 85.1852 | 79.7468 | 78.3951 | 88.6076 | 79.6296 | 82.2785 | 74.6835 | 74.6854 |
| **FDR** | 32.8767 | 35.5556 | 38.8889 | 26.4706 | 37.9310 | 33.3333 | 40.8163 | 40.8980 |
| **FPR** | 14.8148 | 20.2532 | 21.6049 | 11.3924 | 20.3704 | 17.7215 | 25.3165 | 25.3165 |
| **F1** | 65.9933 | 70.7317 | 66.4653 | 70.4225 | 66.4615 | 70.8861 | 67.4419 | 67.4621 |
| **Brier score** | 0.1609 | 0.1514 | 0.1661 | 0.1557 | 0.1531 | 0.1503 | 0.1596 | 0.1597 |

